# Genome-wide Machine Learning Analysis of Anosmia and Ageusia with COVID-19

**DOI:** 10.1101/2024.12.04.24318493

**Authors:** Lucas Pietan, Elizabeth Phillippi, Marcelo Melo, Hatem El-Shanti, Brian J Smith, Benjamin Darbro, Terry Braun, Thomas Casavant

## Abstract

The COVID-19 pandemic has caused substantial worldwide disruptions in health, economy, and society, manifesting symptoms such as loss of smell (anosmia) and loss of taste (ageusia), that can result in prolonged sensory impairment. Establishing the host genetic etiology of anosmia and ageusia in COVID-19 will aid in the overall understanding of the sensorineural aspect of the disease and contribute to possible treatments or cures. By using human genome sequencing data from the University of Iowa (UI) COVID-19 cohort (N=187) and the National Institute of Health All of Us (AoU) Research Program COVID-19 cohort (N=947), we investigated the genetics of anosmia and/or ageusia by employing feature selection techniques to construct a novel variant and gene prioritization pipeline, utilizing machine learning methods for the classification of patients. Models were assessed using a permutation-based variable importance (PVI) strategy for final prioritization of candidate variants and genes. The highest held-out test set area under the receiver operating characteristic (AUROC) curve for models and datasets from the UI cohort was 0.735 and 0.798 for the variant and gene analysis respectively and for the AoU cohort was 0.687 for the variant analysis. Our analysis prioritized several novel and known candidate host genetic factors involved in immune response, neuronal signaling, and calcium signaling supporting previously proposed hypotheses for anosmia/ageusia in COVID-19.

## INTRODUCTION

The COVID-19 pandemic caused by the coronavirus SARS-CoV-2 has had a profound impact on the global population, causing unprecedented health, economic, and social disruptions (Nicola et al. 2020; McKibbin and Fernando 2023). As of May 2024, the World Health Organization (WHO) reported over 775 million confirmed cases and more than 7 million deaths worldwide with over 1 million deaths in the United States (World Health Organization 2024; Centers for Disease Control and Prevention 2024). COVID-19 presents with a variety of symptoms, ranging from mild respiratory issues to severe pneumonia and multi-organ failure (Yuki et al. 2020). Among the diverse manifestations of COVID-19, the loss of smell (anosmia) and loss of taste (ageusia) are notable symptoms, affecting around 50% of infected individuals in western countries (Butowt and von Bartheld 2021). While many recover these senses within weeks, a considerable number experience prolonged or even permanent deficits, which can significantly impact quality of life (Krishnakumar et al. 2023).

Multiple biological hypotheses have been proposed to explain the causes of anosmia and ageusia in COVID-19 patients. The most probable cause of the loss of taste with COVID-19 is the viral invasion of gustatory nerves, taste receptor cells, and the olfactory epithelium. Other proposed contributing factors include an increase in pro-inflammatory cytokines leading to damage of associated cells and elevated or decreased levels of gene and protein factors, such as angiotensin II (Mahmoud et al. 2021; Krishnakumar et al. 2023). For the loss of smell, a primary hypothesis is the death of support cells for olfactory receptor neurons, affecting neuronal function by altering the olfactory epithelium mucus and disrupting the olfactory cilia (Butowt et al. 2023; Anastassopoulou et al. 2024). Another well-supported hypothesis proposes that the host immune response impacts olfactory receptor neuron function by downregulating gene expression for receptors and signaling molecules, alongside inflammation and damage to the olfactory epithelium and olfactory receptor neurons through cytokine activity (Butowt et al. 2023; Finlay et al. 2022). Most proposed biological causes of anosmia and ageusia in COVID-19 are not mutually exclusive and the overall etiology is most likely a combination of factors. The difference between acute or long-term anosmia and ageusia symptoms have also been investigated and may have shared or distinct mechanisms (Anastassopoulou et al. 2024).

Regarding the molecular mechanisms and specific genes and proteins involved in anosmia and ageusia in COVID-19, immune factors such as Interleukin-6 have been implicated (Cazzolla et al. 2020). The largest genome-wide study to date identified 28 variants within the *UGT2A1/UGT2A2* locus significantly associated with loss of smell or taste in COVID-19 (Shelton et al. 2022). However, the genetic mechanisms underlying COVID-19-related anosmia and ageusia have yet to be completely explained.

Machine learning (ML) methods have become widely applied in healthcare research, contributing significantly to our understanding of numerous diseases across various disciplines (Ahsan et al. 2022). ML models offer the advantage of simultaneously evaluating multiple features, identifying patterns and interactions within the data undetected by human observation or traditional analytical methods, making them particularly suitable for analyzing genomic data. Machine learning methods have been applied to several areas within genetics, including transcriptomics, epigenomics, and variant discovery (Reel et al. 2021; Arslan et al. 2021; Ahmed et al. 2021). Within common, complex trait genomic analyses, the traditional approach involves conducting a genome-wide association study (GWAS) using linear or logistic regression to identify individually associated variants with the phenotype. Significant variants identified in the GWAS are then combined to develop polygenic risk scores (PRS). ML methods have been utilized within the GWAS and PRS analyses to perform a multivariate assessment, increase power of the analysis, and prioritize the significant findings (Lello et al. 2019; Ho et al. 2019; Nicholls et al. 2020). ML methods have also been applied with whole genome/exome sequencing (WGS/ES) and genotyping data, where genomic variants are initially filtered based on pathways or previously associated regions, or used to create gene-region features (Behravan et al. 2018; Liu et al. 2022).

Feature selection strategies and pipelines have been applied with success to whole genome/exome sequencing data with ML methods to find and prioritize variants and genomic features (Vadapalli et al. 2022). Classical statistical filtering strategies, such as chi-squared test filters, have been employed for feature selection with model training and classifying Crohn Disease (Romagnoni et al. 2019). Additionally, the Conditional Mutual Information Maximization (CMI) method has been used for selecting features for classifying several diseases (Alzubi et al. 2017). Variable or feature importance analysis is a critical aspect of ML, particularly in medical research and genetics, where the potential for researchers to gain the most insight lies in understanding the utilization of features within the models (Goldstein et al. 2011). When performed correctly, feature selection and model evaluation can produce unbiased results. Permutation-based variable importance (PVI) or feature importance assesses the contribution of each feature in a predictive model and can be assessed across ML models, identifying the features that are significantly influencing predictive performance and are contributing the most important information (Musolf et al. 2022; Fisher et al. 2019).

In this study, we assessed two genome sequencing cohorts for variants and genes associated with the development of anosmia and/or ageusia symptoms in COVID-19. We examined novel feature selection pipelines to construct datasets for ML model to be trained and then tested on held-out test sets to prevent overfitting. A variable importance analysis was leveraged to enhance the interpretability of the models and prioritize the most relevant genetic factors. With this strategy, we found association of several novel genetic factors as well as several known factors including variants and genes involved in immune response, neuronal signaling, and calcium signaling that align with well supported hypothesis in the field.

## RESULTS

The University of Iowa (UI) cohort included a total of 187 individuals, with 9 reporting only a loss of smell, 8 reporting only a loss of taste, and 87 reporting a loss in smell and taste. In total, we had 96 cases and 91 controls for the loss of smell phenotype, 95 cases and 92 controls for the loss of taste phenotype, and 104 cases and 83 controls for the loss of smell and/or loss of taste phenotype (Supplementary Table S1). Statistics for all other symptoms in the cohort were consistent with previously reported COVID-19 symptoms in the population. The assessment of covariates revealed no association of sex, age, and age-squared with the response variable for any of the phenotypes and were not included in subsequent analyses (Supplementary Table S2).

After quality control (QC) filtering and exclusion of variants with multiple alternative alleles the dataset consisted of 9,598,997 variants (38,395,988 features). Our initial examination was on the loss of smell phenotype, where we developed and optimized the feature selection protocols and pipelines. Preliminary testing of our initial filtering pipeline with Conditional Mutual Information-5 (CMI-5), logistic regression (LR), and decision tree variable importance_1000_1 (DT-VI_1000_1) and five ML models with the loss of smell phenotype yielded 1,324 features after filtering and top model performances with the random forest (RF) model with brier score = 0.252 and the support vector machine with polynomial kernel function (SVM-P) model with 58.2% accuracy and 0.577 area under the receiver operating characteristic (AUROC) curve on the held-out test set (Supplementary Table S3). Additional filtering of variants within non-coding regions of the genome yielded a dataset of 300,945 variants (1,203,782 features). We saw an increase in performance with this dataset using the same initial filtering pipeline, CMI-5, LR, and DT-VI_1000_1, yielding 47 features with top performances on the held-out test set with the Lasso (accuracy = 73.0%, AUROC curve = 0.682) and RF (brier score = 0.236) models (Table 1, Supplementary Table S4). Outside of our initial pipeline to determine the viability of our approach, we extensively examined other feature selection strategies. The top performing variant feature selection pipeline was using the combination of filtering algorithms CMI-20, DT-VI_1000_10 (Figure 1), resulting in 1,015 features (Supplementary Table S5) and the extreme gradient boosted tree (XGBTree) model as the top performing model (accuracy = 75.7% [61.9%, 89.5%], brier score = 0.216 [0.113, 0.336], AUROC curve = 0.735 [0.561, 0.909]) on the held-out test set (Table 1, Supplementary Table S6). The top two most important features from PVI for the XGBTree model are from variants in the *SLC2A11* and *LHCGR* genes (Figure 2A, Table 2, Supplementary Table S7). The top performing pipeline for the gene feature analysis was LR (p-value threshold = 0.05), gene feature transformation with the sample allele frequency correction, CMI-20 (Figure 1). This pipeline resulted in 107 gene features (Supplementary Table S5) and Elastic Net (accuracy = 81.1% [68.5%, 93.7%], AUROC curve = 0.798 [0.643, 0.954]) and SVM-P (brier score = 0.192 [0.119, 0.268], AUROC curve = 0.798 [0.648, 0.948]) as the top performing models on the held out-test set (Table 1, Supplementary Table S8). Top features of importance were from the *SNX29P2* and *CDH22* genes for the Elastic Net and SVM-P models respectively, as well as sharing several top features of importance including features from the *SNX29P2*, *BACE2*, and *SLC16A8* genes (Figure 2B,C, Table 2, Supplementary Table S9). All results for all variant and gene feature selection strategies and pipelines are included in the supplemental documentation (Supplementary Table S10-S11).

**Figure 1.**
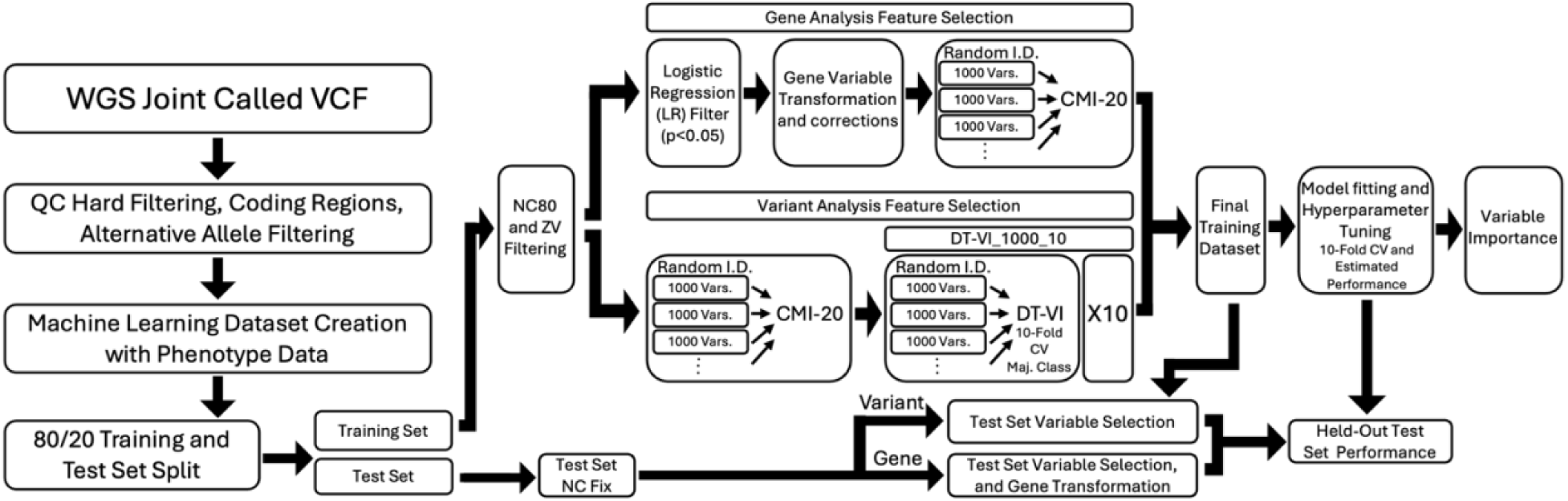
Top performing whole genome sequencing machine learning pipelines for variant and gene analysis. Random I.D. is randomly selected intermediate datasets.

**Table 1.** UI top feature selection pipeline datasets for the loss of smell phenotype held-out test set performance.

| CMI-5, LR, and DT-VI_1000_1 |  |  |  |  |  |  |  |  |  |  |
| --- | --- | --- | --- | --- | --- | --- | --- | --- | --- | --- |
|  | DT | RF | Lasso | NB | SVM-L | SVM-P | SVM-RB | XGBTree | Elastic Net | Log Reg |
| Brier Score | 0.321 | 0.236 | 0.268 | 0.363 | 0.309 | 0.331 | 0.319 | 0.339 | 0.285 | 0.319 |
| Accuracy | 56.8% | 67.6% | 73.0% | 62.2% | 54.1% | 64.9% | 62.2% | 56.8% | 64.9% | 62.2% |
| AUROC Curve | 0.539 | 0.665 | 0.682 | 0.592 | 0.676 | 0.619 | 0.637 | 0.592 | 0.658 | 0.637 |
| CMI-20, DT-VI_1000_10 |  |  |  |  |  |  |  |  |  |  |
|  | DT | RF | Lasso | NB | SVM-L | SVM-P | SVM-RB | XGBTree | Elastic Net | Log Reg |
| Brier Score | 0.293 | 0.223 | 0.273 | 0.323 | 0.274 | 0.247 | 0.256 | 0.216 | 0.279 | 0.256 |
| Accuracy | 51.4% | 67.6% | 64.9% | 67.6% | 56.8% | 67.6% | 43.2% | 75.7% | 64.9% | 43.2% |
| AUROC Curve | 0.503 | 0.731 | 0.679 | 0.628 | 0.622 | 0.685 | 0.313 | 0.735 | 0.655 | 0.313 |
| LR (p-value threshold = 0.05), gene feature transformation with the sample allele frequency correction, CMI-20 |  |  |  |  |  |  |  |  |  |  |
|  | DT | RF | Lasso | NB | SVM-L | SVM-P | SVM-RB | XGBTree | Elastic Net | Log Reg |
| Brier Score | 0.251 | 0.213 | 0.205 | 0.350 | 0.206 | 0.192 | 0.197 | 0.245 | 0.193 | 0.197 |
| Accuracy | 59.5% | 70.3% | 67.6% | 64.9% | 70.3% | 73.0% | 75.7% | 67.6% | 81.1% | 75.7% |
| AUROC Curve | 0.588 | 0.699 | 0.746 | 0.678 | 0.749 | 0.798 | 0.772 | 0.684 | 0.798 | 0.772 |

**Figure 2.**
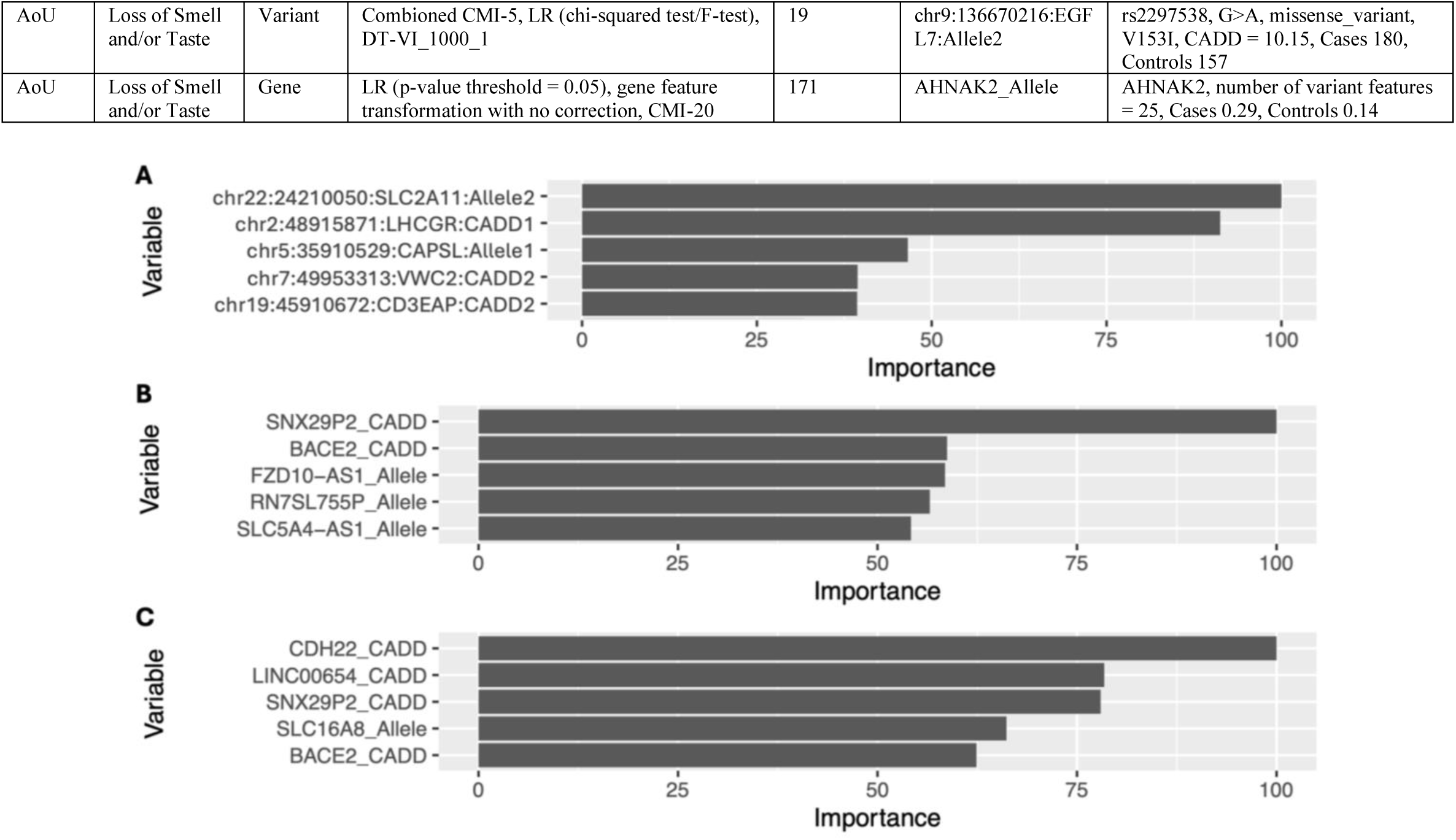
Permutation-based variable importance (PVI) top 5 features for top performing dataset and models for the UI cohort loss of smell analysis. All features included in the model are assessed. (*A*) Variable importance for the extreme gradient boosted tree (XGBTree) model, top performing model for the variant analysis. (*B*) Variable importance for the Elastic Net model, top performing model for the gene analysis according to the accuracy and area under the receiver operating characteristic (AUROC) curve metrics. (*C*) Variable importance for the support vector machine with polynomial kernel function (SVM-P) model, top performing model for the gene analysis according to brier score.

**Table 2.** Summary of machine learning (ML) analysis results for variants and genes in cohort and phenotype top performing datasets. Variant/Gene Information case and control values are counts of samples with the alternative allele for variant features and averages for gene features.

| Cohort | Phenotype | Analysis | Feature Selection Pipeline | Number of Filtered Dataset Features | PVI Top Feature(s) | Variant/Gene Information |
| --- | --- | --- | --- | --- | --- | --- |
| UI | Loss of Smell | Variant | CMI-20, DT-VI_1000_10 | 1,015 | chr22:24210050:SLC2A11:Allele2 | rs4428104, A>C, intron_variant, CADD = 1.466, Cases 74, Controls 42 |
| UI | Loss of Smell | Gene | LR (p-value threshold = 0.05), gene feature transformation with the sample allele frequency correction, CMI-20 | 107 | SNX29P2_CADD | SNX29P2, number of variant features = 3, Cases 3.47, Controls 6.06 |
| UI | Loss of Taste | Variant | CMI-5 | 5,165 | chr10:5037718:AKR1C2:Allele1 | rs78396968, A>G, intron_variant, CADD = 4.417, Cases 16, Controls 19 |
| UI | Loss of Taste | Gene | LR (p-value threshold = 0.05), gene feature transformation with the directional correction, CMI-20 | 103 | GRIN3A_CADD, Y_RNA_Allele | GRIN3A, number of variant features = 4, Cases 35.99, Controls 30.61; Y_RNA, number of variant features = 21, Cases 12.89, Controls 10.87 |
| UI | Loss of Smell and/or Taste | Variant | CMI-5, LR, and DT-VI_1000_1 | 56 | chr11:4621639:TRIM68:CADD2, chr22:39631768:PDGFB:CADD2 | rs2231975, C>T, missense_variant, C442Y, CADD = 0.287, Cases 2, Controls 16; rs17303681, G>T, intron_variant, CADD = 2.866, Cases 4, Controls 11 |
| UI | Loss of Smell and/or Taste | Gene | LR (p-value threshold = 0.05), gene feature transformation with the sample frequency correction, CMI-20 pipeline | 102 | GNAO1_CADD | GNAO1, number of variant features = 8, Cases 5.44, Controls 2.54 |
| AoU | Loss of Smell and/or Taste | Variant | Combiomed CMI-5, LR (chi-squared test/F-test), DT-VI_1000_1 | 19 | chr9:136670216:EGF L7:Allele2 | rs2297538, G>A, missense_variant, V153I, CADD = 10.15, Cases 180, Controls 157 |
| AoU | Loss of Smell and/or Taste | Gene | LR (p-value threshold = 0.05), gene feature transformation with no correction, CMI-20 | 171 | AHNAK2_Allele | AHNAK2, number of variant features = 25, Cases 0.29, Controls 0.14 |

The top performing feature selection pipelines were used to investigate the loss of taste phenotype and the loss of smell and/or loss of taste phenotype. Top results for the loss of taste phenotype for the variant analysis was the single CMI-5 filter and the decision tree (DT) model (accuracy = 62.2% [46.5%, 77.8%], brier score = 0.245 [0.199, 0.301], AUROC curve = 0.647 [0.476, 0.818], Supplementary Table S12) fit to 5,165 features (Supplementary Table S5). PVI for the DT model had the top feature from a variant in the *AKR1C2* gene (Table 2, Supplementary Figure S1A, Supplementary Table S13). For the gene analysis, the top pipeline was LR (p-value threshold = 0.05), gene feature transformation with the directional correction, CMI-20, yielding 103 candidate features (Supplementary Table S5). Top models on the held-out test set were the support vector machine with linear kernel function (SVM-L) (accuracy = 73.0% [58.7%, 87.3%], AUROC curve = 0.709 [0.534, 0.884]) and RF models (brier score = 0.248 [0.205, 0.290]) (Supplementary Table S14). The most important feature from the SVM-L model was *GRIN3A* gene feature and from the RF model was from the Y RNA gene locus. The *snoU13* gene feature was also shared in top features in both models (Table 2, Supplementary Figure S1B,C, Supplementary Table S15). Model performances for all filtering strategies and datasets for the loss of taste phenotype are included in the supplemental documentation (Supplementary Table S16).

As for the loss of smell and/or loss of taste phenotype, the top variant pipeline was CMI-5, LR, and DT-VI_1000_1, filtering to 56 features (Supplementary Table S5) with top model performance on the held-out test set with the RF (accuracy = 64.9% [49.5%, 80.2%], brier score = 0.242 [0.196, 0.290]) and Naïve Bayes (NB) (accuracy = 64.9% [49.5%, 80.2%], AUROC curve = 0.670 [0.487, 0.852]) models (Supplementary Table S17). PVI showed the most important feature for the RF model to be from the *TRIM68* gene and for the NB to be from the *PDGFB* gene. There was also a shared top important variant feature from the *IL21-AS1* gene for the RF and NB models (Table 2, Supplementary Figure S2A,B, Supplementary Table S18). The top gene pipeline was the LR (p-value threshold = 0.05), gene feature transformation with the sample frequency correction, CMI-20 pipeline, selecting 102 gene features (Supplementary Table S5). The top model performance on the held-out test set was with the XGBTree model (accuracy = 73.0% [58.7%, 87.3%], brier score = 0.206 [0.124, 0.297], AUROC curve = 0.778 [0.616, 0.939], Supplementary Table S19). Top feature for the XGBTree model was a *GNAO1* gene feature (Table 2, Supplementary Figure S2C, Supplementary Table S20). Model performances for all filtering strategies and datasets for the loss of smell and/or taste phenotype are included in the supplemental documentation (Supplementary Table S21).

Comparing the filtered datasets from the top performing pipelines for each phenotype we can assess possible shared genetics between the loss of smell and loss of taste with COVID-19. For overlap of variant features, 73 features were intersected with two of the three datasets for each phenotype and three features were found in all three datasets (chr12:131440935:GPR133:CADD1, chr19:56466227:NLRP8:CADD2, chr20:61472938:TCFL5:CADD2). Breaking the features down by position in the genome, two of the three datasets shared 96 features within the same position in the genome and three feature positions agreed across all three datasets (chr12:131440935, chr19:56466227, chr20:61472938). There were 864 genes intersected with features between two datasets and 6 genes (*C1orf127, DOCK1, GPR133, NLRP8, TCFL5, TRIM68*) by features of all three datasets. Comparing the intersection of datasets from the gene analysis, 24 features and 28 genes were overlapped in two of the three datasets and the same four features (BCAT1_Allele, CTC-340A15.2_Allele, MSRA_CADD, PARVB_Allele) and four genes were shared by all three datasets (Supplementary Table S5 and S22).

To examine the generalizability of the genetic association found in the UI cohort with loss of smell and loss of taste symptoms, we sought to test the models and selected genomic features on an independent dataset from the All of Us (AoU) Research Program. The AoU cohort included a total of 947 individuals, with 421 reporting a loss of smell or taste with COVID-19. In total, we had 421 cases and 526 controls for our analysis (Supplementary Table S23). Testing results from the six top performing datasets with all models from the UI cohort variant and gene analyses with the three phenotypes showed little shared association between datasets. Model performances on the held-out test sets were inferior or equal to a majority class model (accuracy = 55.5%) across all datasets and models with ranges in accuracies = [44.5%, 55.6%], brier scores = [0.251, 0.550], and AUROC curves = [0.448, 0.539] (Supplementary Table S24).

The validation results led us to examine the compatibility and similarities between the two cohorts with principal component analysis (PCA). PCA and identity-by-state distance calculations showed the two cohorts to have more variation in the data due to cohort classification than to case/control classification (Supplementary Table S25, Supplementary Figure S3A-L). An assessment of covariates with the AoU cohort also revealed an association with sex (p-value = 0.000367), age, and age-squared (p-values < 2e-16) covariates with the response variable. We then performed a complete gene and variant machine learning analysis with our established best performing feature selection pipelines on the AoU cohort. After QC filtering, exclusion of variants with multiple alternative alleles, and filtering of variants within non-coding regions of the genome, the dataset consisted of 1,583,701 variants (6,334,804 features). First, we performed the analysis without covariates and then with covariates, as well as a covariate only model. The best performing feature selection pipelines were the CMI-5, LR (chi-squared test), DT-VI_1000_1 and CMI-5, LR (F-Test), DT-VI_1000_1 yielding datasets of 9 and 10 features, respectively. Model performance was similar for both feature sets with the CMI-5, LR (chi-squared test), DT-VI_1000_1 dataset having an accuracy = 61.9% and the CMI-5, LR (F-Test), DT-VI_1000_1 pipeline with an accuracy = 61.4% on the held-out test sets with a NB model (Table 3, Supplementary Table S26). A combination of the two datasets (19 features) resulted in similar performance across models (Table 3, Supplementary Table S26). Addition of the covariates to all datasets tested led to the overall best performance with the combined dataset with the top models being Lasso (accuracy = 68.8% [62.2%, 75.4%]), Elastic Net (accuracy = 68.8% [62.2%, 75.4%]), RF (brier score = 0.222 [0.193, 0.251]), and SVM-RB (brier score = 0.222 [0.193, 0.252], AUROC curve = 0.687 [0.609, 0.766]) (Table 3, Supplementary Table S26). PVI showed a variant feature from the *EGFL7* gene to be the most important feature across top models as well as the Age and Age-squared covariates impacting model performances (Figure 3A,B,C, Table 2, Supplementary Table S27). Another notable top feature of importance across models is a variant from the *GRM1* gene. The covariate only models demonstrated slightly stronger performance when considering the metrics brier score and AUROC curve for the held-out test set but weaker performance when considering mean metrics for the 10-fold CV training performance and held-out test set accuracy when compared to the full dataset models (Table 3, Supplementary Table S26). For the gene analysis, the LR (p-value threshold = 0.05), gene feature transformation with no correction, CMI-20, selecting 171 gene features was the top performing pipeline with the RF model (held-out test set accuracy = 65.1% [58.3%, 71.9%], brier score = 0.225 [0.208, 0.241], AUROC curve = 0.678 [0.601, 0.756]) (Supplementary Table S26). The most important gene feature for the RF model from PVI was from the *AHNAK2* gene (Table 2, Supplementary Table S28, Supplementary Figure S4). Model performances for all filtering strategies and datasets for the loss of smell and/or taste phenotype within the AoU cohort is included in the supplemental documentation (Supplementary Table S26).

**Table 3.** AoU top feature selection pipeline datasets for the loss of smell and/or taste phenotype held-out test set performance.

| CMI-5, LR (chi-squared test), DT-VI_1000_1 (9 features) |  |  |  |  |  |  |  |  |
| --- | --- | --- | --- | --- | --- | --- | --- | --- |
|  | DT | RF | Lasso | NB | SVM-L | SVM-P | SVM-RB | Elastic Net |
| Brier Score | 0.276 | 0.328 | 0.253 | 0.302 | 0.253 | 0.272 | 0.274 | 0.253 |
| Accuracy | 56.1% | 55.0% | 57.1% | 61.9% | 51.9% | 54.5% | 55.6% | 57.1% |
| AUROC Curve | 0.534 | 0.534 | 0.571 | 0.574 | 0.560 | 0.524 | 0.563 | 0.571 |
| CMI-5, LR (F-Test), DT-VI_1000_1 (10 features) |  |  |  |  |  |  |  |  |
|  | DT | RF | Lasso | NB | SVM-L | SVM-P | SVM-RB | Elastic Net |
| Brier Score | 0.259 | 0.277 | 0.255 | 0.385 | 0.256 | 0.256 | 0.251 | 0.254 |
| Accuracy | 56.1% | 59.8% | 53.4% | 61.4% | 57.7% | 57.1% | 57.1% | 53.4% |
| AUROC Curve | 0.510 | 0.544 | 0.554 | 0.549 | 0.540 | 0.536 | 0.522 | 0.554 |
| CMI-5, LR (chi-squared test), DT-VI_1000_1 + CMI-5, LR (F-Test), DT-VI_1000_1 (19 Features) |  |  |  |  |  |  |  |  |
|  | DT | RF | Lasso | NB | SVM-L | SVM-P | SVM-RB | Elastic Net |
| Brier Score | 0.276 | 0.271 | 0.258 | 0.381 | 0.257 | 0.275 | 0.270 | 0.256 |
| Accuracy | 56.1% | 57.1% | 57.1% | 61.9% | 57.7% | 57.7% | 58.2% | 57.7% |
| AUROC Curve | 0.534 | 0.554 | 0.592 | 0.591 | 0.588 | 0.556 | 0.552 | 0.592 |
| CMI-5, LR (chi-squared test), DT-VI_1000_1 + CMI-5, LR (F-Test), DT-VI_1000_1 with Covariates |  |  |  |  |  |  |  |  |
|  | DT | RF | Lasso | NB | SVM-L | SVM-P | SVM-RB | Elastic Net |
| Brier Score | 0.243 | 0.222 | 0.232 | 0.380 | 0.231 | 0.231 | 0.222 | 0.229 |
| Accuracy | 61.9% | 65.6% | 68.8% | 61.9% | 67.2% | 67.7% | 68.3% | 68.8% |
| AUROC Curve | 0.554 | 0.677 | 0.671 | 0.655 | 0.668 | 0.664 | 0.687 | 0.671 |
| Covariates Only |  |  |  |  |  |  |  |  |
|  | DT | RF | Lasso | NB | SVM-L | SVM-P | SVM-RB | Elastic Net |
| Brier Score | 0.223 | 0.289 | 0.213 | 0.218 | 0.212 | 0.212 | 0.211 | 0.212 |
| Accuracy | 61.9% | 59.8% | 61.4% | 62.4% | 61.4% | 61.4% | 61.4% | 61.4% |
| AUROC Curve | 0.649 | 0.593 | 0.708 | 0.710 | 0.708 | 0.707 | 0.708 | 0.710 |

**Figure 3.**
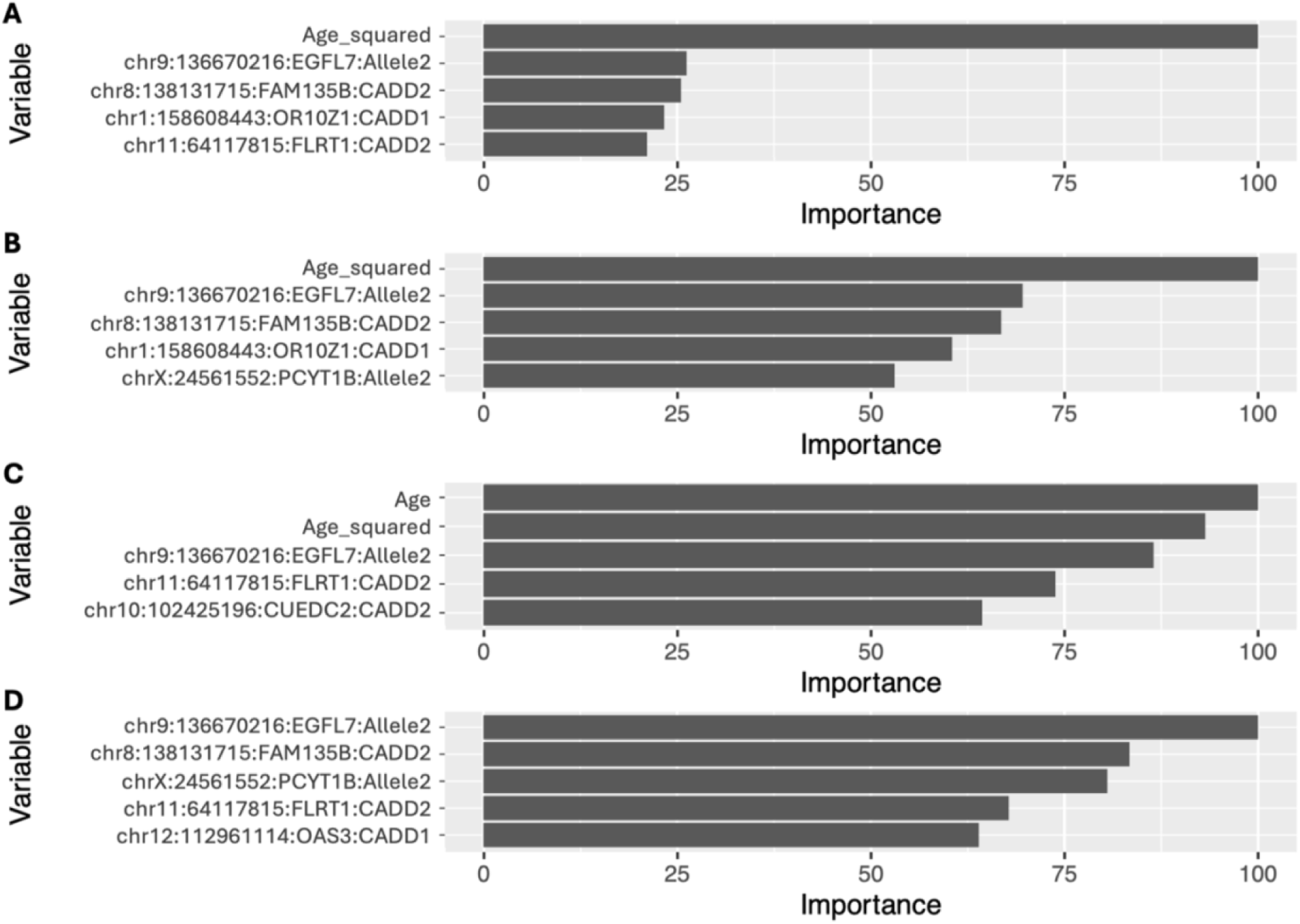
Permutation-based variable importance top 5 features for top performing dataset and models for the AoU cohort variant analysis. All features included in the model are assessed. (*A*) Variable importance for the Lasso model, top performing model for the variant analysis according to the accuracy metric. (*B*) Variable importance for the Elastic Net model (accuracy). (*C*) Variable importance for the RF model (brier score). (*D*) Variable importance for the SVM-RB model (brier score and AUROC curve).

In turn, we extracted the top performing AoU cohort variant dataset features from the UI cohort to validate on an independent dataset. We match the phenotype of loss of smell and/or taste for both cohorts. The validation results were similar to the results of the validation in the reverse direction, with DT (brier score = 0.266), NB (AUROC curve = 0.570), and support vector machine with radial basis kernel function (SVM-RB) (accuracy = 53.3%) models having the strongest held-out test set performance (Supplementary Table S29). Comparing the AoU top variant dataset with the top datasets from the UI cohort analysis, there were no intersection of features or variant positions between the datasets. There was an intersection of features associated with the same genes, with the AoU dataset intersecting the *EGFL7*, *FLRT1*, and *OAS*3 genes with the UI loss of taste variant dataset and intersecting the *LILRB1* gene with the UI loss of smell dataset (Supplementary Table S22).

We tested the 28 variants previously found to be significantly associated with the loss of smell or taste symptoms with COVID-19 for association in the UI cohort and the AoU cohort (Shelton 2022). In the UI cohort, 27 of the 28 variants were present and with all statistical association test performed, none of the variants or variant features were significantly associated with the loss of smell and/or taste phenotype response variable (Supplementary Table S30). Within the AoU cohort, no variants or variant features were significantly associated with the loss of smell and/or taste phenotype (Supplementary Table S30).

The top performing datasets for the variant and gene ML analyses for the UI cohort and AoU cohort were examined with Ingenuity Pathway Analysis (IPA). Individual dataset analysis resulted with 3 datasets with significant overlap of pathways, the UI loss of taste variant analysis dataset, the UI loss of taste and/smell gene analysis dataset, and AoU gene analysis dataset with 72, 2, and 8 significant pathways, respectively (Supplementary Table S31). We found trends within the comparison analysis results showing top overlapping enriched pathways involved in similar biological processes such as immune response, neuronal signaling, and calcium signaling (Supplementary Table S32). The majority of the top pathways have overlap with at least 6 of the 8 datasets (Supplementary Table S32). The upstream regulator analysis found 3 molecules overlapping at least four of the datasets including both cohorts (Supplementary Table S33). Canonical pathway hierarchical clustering resulted in the AoU variant analysis dataset and the UI loss of smell and/or taste variant analysis dataset to be clustered, being the most similar, followed by UI loss of smell variant analysis dataset similar to both datasets. This demonstrates similarities across cohorts with the top performing datasets for each cohort.

## DISCUSSION

In this study, we investigated the host genetic variants and genes associated with anosmia and ageusia symptoms of COVID-19 in two separate cohorts with multiple feature selection strategies and ML methods. We successfully developed novel feature selection pipelines for variant and gene analysis using machine learning models. These models accurately classified patients with and without anosmia and/or ageusia symptoms, significantly outperforming a majority class model. To our knowledge, this is the first study to utilize whole genome sequencing data and ML techniques for gene and variant candidate prioritization and association with anosmia and ageusia with COVID-19. This study improves our understanding of SARS-CoV-2 infections and COVID-19, and specifically the anosmia and ageusia symptoms that impact the daily lives of individuals afflicted by them.

Our genome sequencing analysis included rare and common variants in a joint analysis with the rationale that all variants have an impact on overall biological mechanisms. Including variants with higher and lower allele frequencies in the population would provide a more comprehensive model and analysis. Several studies have used ML for rare variants association with diseases and conditions such as schizophrenia and ovarian failure (Trakadis et al. 2019; Sardaar et al. 2020; Henarejos-Castillo et al. 2021; Jin et al. 2020). Symptom development and severity of COVID-19 has been shown to have a genetic component (COVID-19 Host Genetics Initiative 2021; Severe Covid-19 GWAS Group 2020; Augusto et al. 2023). The genetics underlying symptoms such as anosmia and ageusia in COVID-19 may act as genetic modifiers, with an individual’s genetic background influencing their development with an infection. COVID-19 symptoms are also considered to be complex genetic traits (Zguro et al. 2022). Both common and rare variants have been documented as genetic modifiers and to be involved in the development of complex disease traits (Weiner et al. 2023; Sun et al. 2022; Pinnaro et al. 2020; Pinnaro et al. 2023; Mohar et al. 2024). A previous GWAS assessing common polymorphisms identified 28 variants associated with COVID-19-related loss of smell or taste. However, these variants were not validated in a separate set, and we did not find association of the variants in UI or AoU cohorts, nor was a polygenic risk score evaluation performed. (Shelton et al. 2022). Although our cohorts were underpowered for a GWAS, other studies have shown significantly informative results with smaller sample sizes using genomic data analyzed with ML methods. (Vabalas et al. 2019; Vadapalli et al. 2022). Our goal was to utilize dataset agnostic feature selection strategies for non-bias selection of the most informative features and perform a ML analysis. This allowed us to gain insight into the genetic etiology of anosmia and ageusia with COVID-19 and establish an optimized and robust workflow for the potential analysis of other diseases.

Each cohort included cases of individuals reporting loss of smell and/or taste within the estimated population prevalence for western countries (Butowt and von Bartheld 2021), potentially allowing for more generalizable features to be selected and models to be trained. Optimization of the feature selection pipeline was carried out using the UI cohort with the anosmia phenotype. This approach is responsible for the anosmia variant and gene analyses performing best overall. Validation analysis for both cohorts revealed that the top-performing features and models had poor performance and lacked generalizability to samples in the opposite cohort. Plotting the principal components for each cohort together showed more variation due to the differences in the cohorts rather than differences in the cases and controls, explaining the poor validation performance. These results potentially suggest a more complex genetic basis within the population for anosmia and ageusia symptoms with COVID-19. The AoU cohort results may provide insight into the overall complexity of genes and variants associated, while the UI cohort, with its more limited sample selection, was more focused and found a subset of genetic patterns. Differences with the correlated covariates sex and age also demonstrates the variance between cohorts. A lack of association with the 28 variants previously found to be associated with loss of smell or taste with COVID-19 through GWAS (Shelton et al. 2022) supports the notion of high degree of genetic complexity. All models within each cohort, and for each phenotype in the UI cohort analysis, underwent testing on a complete held-out test set, achieving a local validation. Although the same features, variant or gene, did not allow for validation across cohorts, the overlap of gene functions implicated in the analyses along with several of the same enriched pathways demonstrated both cohort analyses were targeting and finding patterns in the same biological processes.

The top candidate genes from the UI cohort investigating the anosmia phenotype was *SLC2A11* and *SNX29P2*. GLUT11 (*SLC2A11*) has a function of transporting glucose and fructose across the cell membrane and is lowly expressed in most cell types including cells obtained from the nasal cavity and immune cells (Doege et al. 2001; Carithers and Moore 2015; Mick et al. 2020). GLUT11 involvement aligns with a supported hypothesis for the loss of smell involving the decrease in glucose and lack of energy for olfactory cilia (Butowt et al. 2023). The cases/control variant allele counts trends with a decrease in function hypothesis of GLUT11, however, rs4428104 is intron variant with unknown significance. Intron variants may be in an unknown functional element directly influencing gene function or regulation, or they could be in linkage disequilibrium (LD) with the causal variant(s). *SNX29P2* product is hypothesized as a modulator of Sorting Nexin 29 (*SNX29*) with little known about either gene (Sanchez et al. 2020), although SNX29P2 may have a function in immune cells (Orrù et al. 2020).

The top candidate genes from the UI cohort investigating the ageusia phenotype was *AKR1C2*, *GRIN3A*, and Y RNAs. *AKR1C2* encodes an aldo-keto reductase reacting with substrates such as bile, steroids like testosterone, and neurosteroids (Penning et al. 2000; Rižner and Penning 2014). Expression of *AKR1C2* has been found in human tongue tissue and there is evidence of AKR1C2 protein interaction with the SARS-CoV-2 M protein (Zhang et al. 2023; Darapaneni and Jaldani 2021; Samavarchi-Tehrani et al. 2020). There could potentially be a role involving competition of AKR1C2, an unknown substrate, and the SARS-CoV-2 virus in tissue involving taste reception, or with systemic upregulation of SARS-CoV-2 cellular entry factors such as transmembrane serine protease 2 (TMPRSS2) (Mollica et al. 2020).

GRIN3A is a subunit of the N-methyl-D-aspartate (NMDA) receptors and a member of the superfamily of glutamate-regulated ion channels. *GRIN3A* is involved in glutamate neuronal signaling with associated pathways intersecting across cohort and phenotype analyses in the present study. Glutamate has been shown to be the neurotransmitter of olfactory and gustatory signaling and GRIN3A protein function could potentially be involved with the anosmia and/or ageusia symptoms with COVID-19 (Berkowicz et al. 1994; Roper 2013). Y RNAs are conserved non-coding RNAs that appear to have a function with chromosomal DNA replication (Christov et al. 2006). Certain Y RNAs have been found to be associated with severity of symptoms with a viral infection, including COVID-19 (Olliff et al. 2023).

The top candidate genes from the UI cohort investigating the anosmia and/or ageusia phenotype were *TRIM68*, *PDGFB*, and *GNAO1*. The TRIM68 protein functions as E3 ubiquitin ligases and has known functions inside inflammatory pathways. TRIM68 decreases or stops production of interferon with viral detection (Wynne et al. 2014). The missense variant (rs2231975, C442Y) could potentially alter the structure or function of TRIM68 protein in a way that is activated in the presence of a SARS-CoV-2 infection to contribute to or protect from the development of loss of smell and/or taste. Given the amino acid position and TRIM68 protein structure, the loss of cysteine likely would not affect disulfide bonding, while the change to tyrosine may enable post-translational phosphorylation of the tyrosine side chain hydroxyl group. The protein encoded by *PDGFB* plays a role in a wide range of processes including angiogenesis (Guo et al. 2003). A previous study has implicated *PDGFB* with susceptibility and severity of COVID-19 symptoms (Chung et al. 2022) and it has been hypothesized that *PDGFB* could be involved with an enhanced immune response with a SARS-CoV-2 infection (Okamoto and Ichikawa 2021). The case/control alternative allele counts for the associated variants with *TRIM68* (rs2231975) and *PDGFB* (rs17303681) show higher counts in controls than cases. For *TRIM68*, this trend could suggest a protective effect of the variant and for *PDGFB* the trend aligns with a damaging effect, but in this scenario, no development of symptoms. *GNAO1* encodes the alpha subunit of the Go heterotrimeric G-protein signal-transducing complex and has been associated with neurodevelopmental disorders (Yang et al. 2023). The GNAO1 protein is also involved in the release of inflammatory mediators from immune cells (Jin et al. 2016) and has been associated with inhibitory signaling in olfactory transduction in rats (Corey et al. 2021).

The top candidate genes from the AoU cohort investigating the anosmia and/or ageusia phenotype were *EGFL7*, *GRM1*, and *AHNAK2*. *EGFL7* encodes an endothelial cell secreted protein that plays a role in vasculogenesis, angiogenesis (Usuba et al. 2019), and is involved in calcium binding (Davis 2010). In recent studies, EGFL7 was implicated in COVID-19 severity (Iosef et al. 2023) and *EGFL7*-knockout mice displayed dysfunction in olfactory behavior (Bicker et al. 2017). The case/control alternative allele counts for the associated variant (rs2297538, V153I) trends with a decrease in proper function hypothesis of EGFL7. The substitution of a valine to isoleucine is a chemically conservative variant with a solvent-exposed side chain, likely not disrupting the protein structure alone. However, the loss of the valine could potentially affect protein binding or domain-specific functions or interactions. The *GRM1* gene encodes a metabotropic glutamate receptor that is expressed in taste buds and is involved in umami taste perception and included in the canonical pathway sensory perception of taste (Diepeveen et al. 2022). The *AHNAK2* gene encodes a nucleoprotein that is associated with calcium channel proteins (Komuro et al. 2004). In a previous study, a rare variant analysis investigating genetic association with severe COVID-19 identified a strong signal from a variant in the AHNAK2 gene (Matuozzo et al. 2023).

By examining the trends within and across the datasets, phenotypes, and cohorts we can obtain a more general view of host genetics involved with the loss of smell and taste in COVID-19. We had several features, genomic positions, and genes intersect between datasets and phenotypes within the UI cohort, and four genes, *EGFL7*, *FLRT1*, *OAS3*, and *LILRB1* overlap between the two cohorts. *OAS3* and *LILRB1* are both involved in immune response (Zhang et al. 2017; Kristiansen et al. 2011) and *OAS3* has been implicated in COVID-19 susceptibility and severity (Abdelhafez et al. 2023). The IPA comparison analysis of all top performing feature sets highlighted three biological processes, immune response, neuronal signaling, and calcium signaling, that are in line with the implicated genes described above.

Overall, the results of this study provide additional evidence for previous hypotheses proposed for the loss of smell and/or taste with COVID-19 (Mahmoud et al. 2021; Krishnakumar et al. 2023; Butowt et al. 2023; Anastassopoulou et al. 2024; Finlay et al. 2022). Our results provide support for the potential compound involvement of multiple processes. Starting with the increase in vascularization and/or decrease in vascular integrity (*EGFL7*) to the olfactory epithelium and gustatory nerves, allowing for an increased immune and inflammatory response assisted by other factors such as *SNX29P2*, *TRIM68*, *PDGFB*, *GNAO1*, *OAS3,* and *LILRB1*. The increased inflammation potentially causes damage to the olfactory epithelium, olfactory receptor neurons, and gustatory nerves, and is accompanied by an increase in viral invasion (*AKR1C2*) of gustatory nerves and the olfactory epithelium including the support cells. Damage of the support cells alters the olfactory epithelium mucus contributing to the decrease in energy for olfactory cilia function (*SLC2A11*). Altogether with the potential alterations in olfactory and gustatory transduction with factors such as *GRIN3A* and *GRM1* glutamate neuronal signaling, *GNAO1* involved signaling, and calcium signaling (*GRIN3A, PDGFB*, *GNAO1, AHNAK2, EGFL7*, *GRM1*). These processes, altogether or in subsets, may cause the loss of smell and/or taste with COVID-19 (Figure 4).

**Figure 4.**
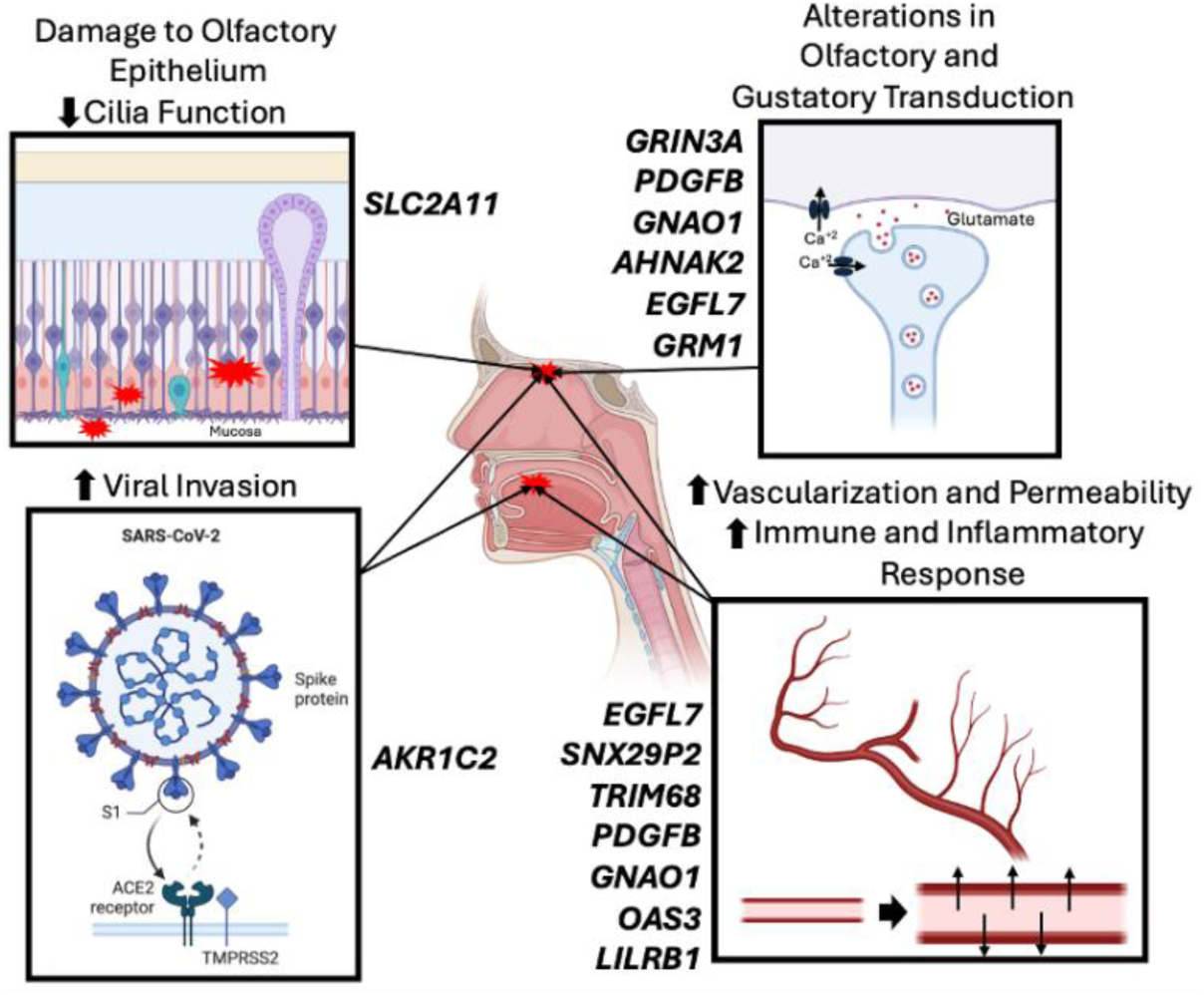
Summary of hypothesis for the involvement of prioritized candidate genes and biological processes in the development of anosmia and ageusia with COVID-19. Created in BioRender. Pietan, L. (2025) https://BioRender.com/h04s965

A main strength of this study is the robust ML pipeline and analysis. With whole genome sequencing data, we examined numerous feature selection strategies to cut down on the high dimensional nature of the data in a performance enhancing way. We chose not to introduce dataset specific biological filters to increase our chances for novel findings. Our established pipelines were optimized on the UI anosmia phenotype dataset. When validating on other datasets, variations of the pipeline were able to achieve insightful performance demonstrating that these feature selection algorithms, individually and in combination, can provide informative results with ML models. The variations of the pipeline should be tested with new datasets in search of optimal performance. In line with best practices, we implemented the 80/20 dataset split, to create a training/validation set and a held-out test set. The split was performed before feature selection, eliminating any biases and data leakage. Hyperparameter tuning of the ML models was implemented inside a stratified 10-fold CV, altogether estimating performance on validation sets and assessing true predictive performance on a held-out test set within cohorts. Model selection with 10-fold CV was based on the brier score. Model selection based on other metrics yielded weaker results when assessed on the held-out test set. While assessing overall pipeline and model performance, both training and test set performance were considered, with priority given to the brier score, accuracy, and AUROC curve metrics. Multiple well-established and validated ML models were tested and had previously been demonstrated to produce insightful results on biological datasets (Ahsan et al. 2022). With our whole genome sequencing top performing datasets and pipelines, tree-based models, SVM models, Lasso, and Elastic Net were consistently at the top for test set performance and demonstrated less overfitting. With all datasets and pipelines tested for the UI anosmia phenotype, the SVM-RB model demonstrated slightly less consistency, notably with gene analysis pipelines. The DT model does have weaker performance in general but when paired with a DT-VI filter it had a more notable performance with several datasets. Lasso and Elastic Net have the advantage of being interpretable as logistic regression models and DT models can be visualized for interpretation. To conclude the pipeline, PVI assessment for each model was completed and is an important element for the overall goal of the study. Resulting PVI metrics can be compared across models and are not model specific. Ultimately, PVI, feature selection, and pathway analysis allowed for genomic factor prioritization.

The present study is not without limitations. One of the main limitations is the sample size of the UI cohort and incomplete validation of results on an independent dataset. With limited samples sizes, the features selected, and models trained may not be generalizable to the overall population and only target a subset of the population. Future studies aim to validate the results obtained from the UI and AoU cohorts in other datasets. Another limitation is with patient self-reporting of their COVID-19 symptoms and through surveys with the UI and AoU cohort. Asking patients in person or through surveys to assess their own symptoms without clear definition (metric-based), potentially with significant time after symptoms subsided, leads to inconsistencies between cases and controls adding noise to the analysis. The final main limitation with the analysis is not having the ability to control for SARS-CoV-2 infection strain or including a temporal and geographical (AoU cohort) assessment of cases and controls as proxy. Previous studies have shown differences in COVID-19 symptoms with SARS-CoV-2 strains (Kumar et al. 2021, Miwa 2023, Anastassopoulou 2024). For future studies we plan to further validate and assess our variant and gene ML pipeline analysis on additional disease datasets.

In conclusion, we sought to prioritize candidate variants and genes involved in the development of anosmia and ageusia with COVID-19 using a novel pipeline of feature selection techniques, ML methods, and a variable importance assessment. We examined two separate cohorts and prioritized variants and genes in overlapping biological processes including immune response, neuronal signaling, and calcium signaling. These findings align with existing literature and offer additional support to previously proposed hypothesis in the field for the development of anosmia and/or ageusia with COVID-19.

## METHODS

### University of Iowa COVID-19 Whole Genome Sequencing Cohort (UI cohort)

Patients were recruited and the participant or their lawful authorized representative signed an informed consent following approval from the Iowa Institutional Review Board (University of Iowa IRB protocol # 202005052). In total, 187 patients participated, all reporting a positive SARS-CoV-2 infection with antibody or polymerase chain reaction (PCR) testing performed at the University of Iowa Hospital and Clinics and surrounding areas or presumed positive by being symptomatic and in a household with someone who tested positive. Eighteen symptoms were collected from patients through surveys or their electronic medical records including fever, chills, dry cough, productive cough, productive cough with blood, fatigue, muscle or joint pain, sore throat, nausea or vomiting, loss of sense of taste (aguesia), loss of sense of smell (anosmia), rash, runny nose or congestion, headache, shortness of breath, cold sores or sores in the mouth, diarrhea, conjunctival congestion, and no symptoms. The anosmia and/or ageusia phenotypes were constructed from this data and all symptoms were used to assess dataset consistency with population statistics for COVID-19. Loss of smell, loss of taste, and a combined loss of smell and/or loss of taste are the phenotypes that were assessed in the UI cohort with the analysis described below. Patients with COVID-19 and reporting either a loss of smell and/or a loss of taste were labeled as cases and patients with COVID-19 and did not have a loss of smell and/or loss of taste were considered controls. Patients under four years of age at time of consent were excluded from the study due to possible inability to assess and convey symptoms.

Whole blood or saliva was obtained from each patient for DNA extraction and whole genome sequencing. Saliva samples were collected in OGR-600 collection kits (adults) or OC175 collection kits (minors too young to spit, or adults with trouble spitting) (DNA Genotek, Ontario, CAN). Symptom surveys were returned with saliva samples. DNA was extracted using PrepIT reagent and ethanol precipitation per manufacturer’s instructions (DNA Genotek). Blood samples were collected in EDTA tubes from consented patients. Symptom surveys were collected after consent. DNA was extracted using Gentra Puregene Blood Core Kit C according to manufacturer’s instructions (Qiagen, Hilden, Germany). DNA was assessed for purity on a Nanodrop 2000 (260/280 >= 1.8; 260/230 >= 1.0) (Thermo Fisher Scientific, Waltham, Massachusetts, USA). DNA concentration was measured by fluorescence (Qubit 2.0, BR assay), DNA was diluted, checked for fragmentation with 1% agarose gel, and concentration confirmed by HS Qubit assay (Thermo Fisher Scientific). DNA was submitted for whole genome sequencing to Novogene (Sacramento, California, USA) or the Genomics Division of the Iowa Institute of Human Genetics and sequenced using a NovaSeq6000 (Illumina, San Diego, California, USA).

Dataset curation, preprocessing, and machine learning analysis was performed on the University of Iowa’s high-performance computing cluster. Short read whole genome sequencing data was processed using the DRAGEN Bio-IT Platform V3.8 from Illumina. The DRAGEN DNA Pipeline (DRAGEN Host Software Version 07.021.602.3.8.4 and Bio-IT Processor Version 0×18101306) mapped reads to the GRCh37 reference genome and variants were called with default parameters. Sequencing statistics were calculated using MultiQC (Ewels et al. 2016). Individual samples were jointly call and quality control filtering of variants was performed with the Genome Analysis Toolkit (GATK, version 4.1.5.0) using the following thresholds, DP < 8.0 (Depth), QD < 2.0 (QualByDepth), QUAL < 30.0 (Quality), SOR > 3.0 (StrandOddsRatio, FS > 60.0 (FisherStrand), MQ < 40.0 (RMSMappingQuality), MQRankSum < -12.5 (MappingQualityRankSumTest), ReadPosRankSum < -8.0 (ReadPosRankSumTest) (Van der Auwera and O’Connor 2020). The Ensembl Variant Effect predictor (VEP, version 105) was used to annotate variants for gene symbols and Combined Annotation Dependent Depletion (CADD) scores with the pick_allele option set to only annotate one block of consequence data per variant allele (McLaren et al. 2016; Rentzsch et al. 2019). Samples were further assessed for outliers with PLINK v1.9 (www.cog-genomics.org/plink/1.9/, Chang 2015). Sex, age, and age squared were examined as potential covariates to include in models. Chi-squared test and logistic regression was performed to determine any association of covariates with the response variable. No ancestral correction was performed as the majority of individuals included in the study were white, non-Hispanic. However, to confirm no correction for residual population structure was needed, PLINK v1.9 logistic regression association tests with different number of principal components (PCs) were used and visualized with quantile–quantile plot (Q-Q plot). The PCs and results conforming most to ideal and bias controlled conditions were used in subsequent analyses. For the machine learning analysis, additional filtering was performed to exclude variants with multiple alternative alleles and include only variants within coding regions of the genome as annotated by GENCODE v42 with 20 base pairs at each end. The bed file was obtained using the UCSC Table Browser (https://genome.ucsc.edu/cgi-bin/hgTables) and filtering was accomplished using BCFtools (version 1.12) (Frankish et al. 2021; Karolchik et al. 2004; Danecek et al. 2021).

### All of Us Research Program COVID-19 Whole Genome Sequencing Cohort (AoU cohort)

The All of Us Research Program (https://www.researchallofus.org/) participant data was obtained and analyzed through the Researcher Workbench. All analyses for the AoU cohort were performed on the Cloud Analysis Terminal powered by the Google Cloud Computing Platform. Through the Cohort Builder, individuals were selected based on availability of short read whole genome sequencing data and their participation and answers in the COVID-19 Participant Experience (COPE) Survey. Participants were included in the present study if they answered “Yes” to the question “Were you tested for COVID-19 in the past month?, “Yes” to the question “Was the test for COVID-19 positive?”, and reported their symptoms to the question “Which of the following symptoms did you have? (select all that apply).” and did not skip. The symptom options available were, “A fever/feverish”, “Chills or shivers (feeling too cold)”, “Unusual fatigue”, “Unusually strong muscle pains/aches”, “Skipping meals”, “Cough”, “Sore or painful throat”, “Difficulty breathing or shortness of breath”, “Unusually hoarse voice”, “Unusual chest pain or tightness in your chest”, “Runny or stuffy nose”, “Loss of smell or taste”, “Unusual eye soreness or discomfort (e.g. light sensitivity or excessive tears)”, “Raised, red, itchy, welts on the skin or sudden swelling of the face or lips”, “Headache”, “Dizziness or light-headedness”, “Confusion, disorientation, or drowsiness”, “Unusual abdominal pain or stomach ache”, “Diarrhea”, “Nausea, “Red/purple sores or blisters on your feet, including your toes”, and “None of the above (if selected, no other response options are available.” Only the anosmia and/or ageusia phenotype was available and assessed in the AoU cohort. Participant case/control classification for the anosmia and/or ageusia phenotype was determined by the inclusion or absence of “Loss of smell or taste” in the symptoms reported. All versions of the survey were used, and questions were required to be answered within the same survey attempt based on the survey_datetime. Multiple attempts of the survey by participants were screened to ensure symptoms were consistent across attempts. To control for ancestral background within the dataset, the cohort was filtered to only include participants that were classified as European (eur) according to the All of Us genetic predicted ancestry. Samples were filtered if they were included in the flagged samples documentation determined by the All of Us sample QC pipeline. In total, the cohort consisted of 947 samples. Sex, age, and age squared were examined as potential covariates to include in the analysis. A chi-squared test and logistic regression was performed to determine any association of covariates with the response variable and included if association was found. Short read whole genome sequencing data was extracted from the database using the described cohort. Variants were filtered according to the All of Us variant QC pipeline and recommendations. For the machine learning analysis, additional filtering was performed to exclude variants with multiple alternative alleles and include only variants within exon regions of the Gencode v42 basic transcripts provided by the All of Us exome file. Filtering was accomplished using BCFtools (version 1.12) (Frankish et al. 2021; Danecek et al. 2021). Variants were annotated for gene symbols and Combined Annotation Dependent Depletion (CADD) scores with the pick_allele option set to only annotate one block of consequence data per variant allele with VEP (version 105) (McLaren et al. 2016; Rentzsch et al. 2019).

### Cohort Comparison

VCF files from the UI cohort and the AoU cohort were used to perform a principal component analysis (PCA) using PLINK v1.9 to compare the essential features of the two datasets and assess similarity (Chang et al. 2015). Individual cohorts were assessed with cases and controls (loss of smell or taste), and several comparisons were made between cohorts with and without case and control annotations. PCA was performed with QC filtered whole genome sets, whole genome sets with intersected variants, variant sets after coding regions and multiple alternative allele filtering (ML dataset), and the intersected variant sets after coding regions and multiple alternative allele filtering. Each comparison was quantified using a permutation test (10,000 permutations), permuting group labels for between group identity-by-state (IBS) differences, where the groups are either cases and controls or cohort designations.

### Machine Learning Dataset Creation

The machine learning analysis was performed in the R (version 4.3.1) and Python (version 3.10.12) programming languages, using R, Python, and Linux command-line libraries, packages, and tools (R Core Team 2022, Python Software Foundation 2023). All random seeds were set to seed = 123. The input dataset to the machine learning analysis was created from all variants that passed filtering described above. The python package, VCF Parser, was used to parse the VCF files and create all features (predictor variables) (Magnusson 2016). Four features were generated for each variant across samples: a pair of allele features representing the first and second alleles (encoded as 0 for reference allele and 1 for alternative allele), along with corresponding CADD features with the reference allele encoded as 0 and the alternative allele encoded with the CADD score. The feature names were formatted as chr:pos:gene symbol:feature type, with “chr” and “pos” being the chromosome number and position in the genome of the variant and “feature type” being the first or second allele or CADD feature (e.g. chr9:136670216:EGFL7:Allele2). The target variable or response variable for each dataset was coded as a 1 or 0 for cases and controls, respectively. To address multiple model testing and provide a measure against overfitting and detection of false positive patterns, we performed a random 80/20 split of the full dataset, where 80% of the samples were included in a training set and 20% of the samples reserved for a held-out test set. Features were removed from the training set if 80% or more of the samples recorded a no call for a feature (i.e. “.” recorded for alleles in the VCF file) (NC80 filtering). If the feature is below the 80% threshold, no calls were recorded as 0 for the reference allele. If a feature had zero variance among training samples, it was removed from the training set (ZV filtering). We extensively examined different feature selection strategies with variant features and gene features before model training. We selected the top performing strategies based on held-out test set performance metrics of models trained on the filtered features. We tested filters based on mutual information, logistic regression, and model-based features importance. Components of the top performing filtering pipelines are described in the following section. A comprehensive list of filtering strategies and methods tested are described in the supplemental methods (Supplementary Materials and Methods S1).

### Conditional Mutual Information Maximization Filtering (CMI)

The input dataset was partitioned into 1000 randomly sampled features without replacement for filtering. The intermediate datasets were input to a conditional mutual information maximization filtering algorithm performed with the Praznik (version 11.0.0) and Recipes (version 1.0.10) R packages (Kursa 2021; Kuhn et al. 2024). This algorithm selects the feature from the dataset with the highest mutual information and selects subsequent features based on the highest information gain given the already selected features. For this study, the number of selected features at each intermediate dataset was 5 features (CMI-5) and 20 features (CMI-20). All features selected from intermediate dataset filtering were included in downstream analysis.

### Logistic Regression Filtering (LR)

All input features from a dataset are assessed individually with the response variable with logistic regression. P-values were calculated either with a chi-squared test or F-test. All features with p-values exceeding a set threshold are included in downstream analysis. The thresholds used with the top performing pipelines were p-value < 0.10 and p-value < 0.05.

### Decision Tree Variable Importance Filtering (DT-VI)

Input datasets were randomly sampled, selecting 1000 features at a time, without replacement, to construct intermediate datasets similar to the conditional mutual information maximization filtering. This method was tested with and without the allele features converted to a factor. Decision tree models were trained on the intermediate datasets for classification of the response variable using the MachineShop R package (version 3.7.0) (Smith 2024). Decision Tree model hyperparameter tuning was completed using a grid search with stratified 10-fold cross-validation (CV) for model and parameter selection. Parameter values tested for the decision tree model are listed in supplementary methods (Supplementary Table S34). Estimated predictive performance from the 10-fold CV was recorded as mean accuracy. If the best performing decision tree model for an intermediate dataset exceeded the accuracy threshold of a majority class model for the training set, relative permutation-based variable importance (PVI, samples = 25) was performed. All features with a variable importance score > 0 passed the filter and were included in the downstream analysis. Intermediate datasets were either constructed once for filtering, as a filtering step in a feature selection pipeline, or a total of ten iterations were performed with different samplings each iteration. The notation for these algorithms that will be reference throughout are DT-VI_X_Y, with the X being the number of features randomly sampled for intermediate datasets and the Y being the total number of iterations through the algorithm (e.g. DT-VI_1000_1 and DT-VI_1000_10, respectively mentioned in this section)

### Gene Feature Transformation and Analysis

In our gene feature analysis, filtered and non-filtered variant features from the training set were input to a gene feature transformation. The transformation consisted of binning variant features by gene symbol annotation and using an additive approach, summing the features within a bin. All allele features were summed, and all CADD features were summed within a gene, creating two gene features for each gene if features were included in the input dataset. Features not annotated with a gene symbol, were binned in 25 kb windows across the genome, generating region features. The naming convention of these features is gene symbol underscore feature type, with feature type either “Allele” or “CADD” (e.g. SNX29P2_CADD). Three transformations were tested including a no correction transformation, a sample allele frequency correction transformation, and a directional correction transformation. The no correction transformation has no correction and transforms the features as is, directly from the input dataset. The sample frequency correction transformation assesses the frequency of the alternative allele in the training samples. Alternative allele frequencies greater than 0.5 in the training set had the feature encoding swapped, encoding reference alleles as 1 (or their CADD score for CADD features) and alternative alleles as 0. The directional correction transformation assesses each feature and attempts to correct for directional effects. This is done by assessing the ratio of alternative alleles corresponding to cases. If alternative alleles are present more often in controls than in cases or the ratio metric is less than the threshold set at 0.5, the encoding of the feature is swapped. The gene features were then used in downstream analysis either with filtering of the gene features or input to ML model training.

### Feature Selection Pipelines

Feature (variable) selection (filtering) pipelines will be referenced throughout with the notation X, Y, Z. The X filtering method is performed first with the output of X used for the input to the Y feature selection method, and if applicable, followed by Z. The output dataset of Z is the input dataset to the ML analysis, model training and testing on the held-out test set and PVI analysis. All feature selection pipelines, and the ML analysis were optimized on the UI cohort with the anosmia phenotype. Our initial filtering strategy was CMI-5, LR (p-value threshold = 0.1), DT-VI_1000_1 that we performed across phenotypes and cohorts. Our top performing pipelines were CMI-20, DT-VI_1000_10 for the variant analysis and LR (p-value threshold = 0.05), gene feature transformation (with and without corrections), CMI-20 for the gene analysis, both tested across phenotypes and cohorts.

### Machine Learning Analysis

After feature selection, models were trained on the training sets. The models assessed were decision tree (DT), random forest (RF), extreme gradient boosted tree (XGBTree), Lasso, Elastic Net, logistic regression (log reg), naïve bayes (NB), support vector machine with radial basis kernel function (SVM-RB), support vector machine with linear kernel function (SVM-L), and support vector machine with polynomial kernel function (SVM-P). A grid search with stratified 10-fold cross-validation (CV) was used for hyperparameter tuning and model selection. Hyperparameter values for each model are listed in supplementary methods (Supplementary Table S34). 10-fold CV estimate predictive performance was recorded as mean metrics including brier score, accuracy, Cohen’s kappa, area under the receiver operating characteristic (AUROC) curve, sensitivity, and specificity. Models were selected based on brier score, due to brier score being a proper scoring function, with measuring of the accuracy based on prediction probabilities. Held-out test sets were corrected for no calls and features included in model training were selected. For the gene analysis, gene features were constructed in the test sets by selecting the variant features used for the gene transformations in the training set and accounting for any corrected features, if performed. The models selected were used for classification of samples in the held-out test set and true predictive performance of the models were recorded with the same metrics listed above.

Confidence intervals were calculated at 95% for the top-performing pipelines and models, using the normal approximation method for accuracy, the bootstrap method for brier score, and the DeLong method for the AUROC curve. For each model, relative permutation-based variable importance (PVI, samples = 25) was calculated and reported for each input feature. The variable importance score is based on the mean change in brier score, and scores are scaled to from 0 to 100, with the most important feature having the score of 100. Feature selection, model performance, and PVI were all used for final candidate variant and gene prioritization.

### Validation

The top-performing datasets and models from the UI cohort were tested on the AoU cohort, and similarly, the top-performing dataset and models from the AoU cohort were tested on the UI cohort. For the UI cohort VCF file, variants present in the final datasets were lifted over to the GRCh38 reference genome using the BCFtools plugin liftover (version 1.18) to construct the datasets for training of models for validation on the AoU dataset and for the validation of the AoU cohort models (Genovese et al. 2024). For each of the top performing datasets from each cohort, the training and the testing sets were combined, and models were trained on the complete cohort datasets. Variants were matched with reference and alternative alleles across cohorts and variants that were not present in the validation cohort were recorded as 0/0 or the reference allele across all samples. Variants with multiple alternative alleles were encoded as 0 for the reference allele and 1 for all alternative alleles. Models were then tested on the independent held-out test sets and evaluated for predictive performance.

Within each cohort, we tested association of a 28 variant candidate region, found to be significantly associated with the loss of smell or taste phenotype with COVID-19 in a 23andMe cohort (Shelton et al. 2022). Two sets of features were constructed for the variant set, features with the same encoding strategy as described above for the machine learning analysis with two allele features for each variant and an additive encoding strategy used in the 23andMe cohort genome-wide association study (Shelton et al. 2022). Three statistical tests were performed to test for association of predictor variables with the response variable, chi-squared test, logistic regression with the Wald test, and logistic regression with a likelihood ratio test. The likelihood ratio test was used to test for association in the 23andMe cohort genome-wide association study (Shelton et al. 2022).

### Ingenuity Pathway Analysis (IPA)

The top performing datasets for the machine learning analysis for both cohorts were submitted for pathway analysis with Ingenuity Pathway Analysis (IPA) (QIAGEN Inc., https://www.qiagenbioinformatics.com/products/ingenuity-pathway-analysis) (Kramer et al. 2014). We submitted all genes associated with variant or gene features for a core analysis of individual datasets examining overlap with pathways, upstream elements, and disease and biological functions. With the core analysis, default parameters were used and “Anosmia” and “Ageusia” were added as causal networks to be assessed. Analysis significance was determined by uncorrected and Benjamini-Hochberg multiple test corrected p-values. We then performed a comparison analysis of all core analyses performed to assess overlap between datasets, phenotypes, and cohorts with canonical pathways and upstream regulators. Each dataset and pathway are given a score, the negative log of the p-value derived from the right-tailed Fisher’s Exact Test, which we averaged across datasets to yield a rank by overall score.

## DATA ACCESS

The WGS data generated in this study have been submitted to the NCBI BioProject database (https://www.ncbi.nlm.nih.gov/bioproject/) under accession number PRJNA1173221. Variant data and ML datasets from the UI cohort analyzed during the study are available from the corresponding authors on request. The AoU cohort WGS and ML data was obtained through the All of Us Research Program’s Controlled Tier Dataset v7 available to authorized users on the Researcher Workbench. R libraries used by this study are publicly available at the CRAN website: https://cran.r-project.org (accessed on 25 April 2024). All Python libraries used by this study are publicly available. Software used for the study can be found at the GitHub website: https://github.com/lpietan/COVID-19 (accessed on 20 June 2024).

## ETHICS STATEMENT

The study was conducted in accordance with the Declaration of Helsinki. For the UI cohort, patients presenting to the University of Iowa with COVID-19 were consented for genetic studies approved by the University of Iowa Institutional Review Board (IRB#: 202005052). Informed written consent was provided by all patients in the UI cohort in accordance with a protocol approved by the University of Iowa Institutional Review Board (IRB Number 202005052), and studies were conducted in accordance with the Belmont Report. The Institutional Review Board of the University of Iowa gave ethical approval for this work.

## COMPETING INTEREST STATMENT

The authors declare no conflict of interest.

## Data Availability

All data produced in the present study are available upon reasonable request to the authors.

## ACKNOWLEDGMENTS

This research was funded by the Interdisciplinary Genetics T32 Predoctoral Training Grant, grant number T32 GM 008629, the Integrated DNA Technologies Bioinformatics Fellowship Program and the Stead Family Department of Pediatrics departmental funds. The University of Iowa genome sequencing was funded in part by the Stead Family Department of Pediatrics start-up funds for Hatem El-Shanti.

We gratefully acknowledge the University of Iowa participants and the All of Us participants for their contribution, without whom this research would not have been possible, and we thank the National Institutes of Health’s All of Us Research Program for making available the participant data used in this study. We thank the Shivanand R. Patil Cytogenetics and Molecular Laboratory and the Genomics Division of the Iowa Institute of Human Genetics (IIHG).

## AUTHOR CONTRIBUTIONS

Conceptualization, L.P., B.D., T.B. and T.C.; Methodology, L.P., B.J.S., B.D., T.B. and T.C.; Software, L.P.; Validation, L.P.; Formal Analysis, L.P.; Investigation, L.P.; Resources, H.E. and B.D.; Data Curation, L.P., E.P., M.M., H.E. and B.D.; Writing—Original Draft Preparation, L.P.; Writing-Review and Editing, L.P., E.P., H.E., B.J.S., B.D., T.B. and T.C.; Visualization, L.P.; Supervision, B.D., T.B. and T.C.; Project Administration, B.D., T.B. and T.C.; Funding Acquisition, L.P., H.E., B.D., T.B. and T.C.

## REFERENCES

1. Abdelhafez M, Nasereddin A, Shamma OA, Abed R, Sinnokrot R, Marof O, Heif T, Erekat Z, Al-Jawabreh A, Ereqat S. 2023. Association of IFNAR2 rs2236757 and OAS3 rs10735079 Polymorphisms with Susceptibility to COVID-19 Infection and Severity in Palestine. Interdiscip Perspect Infect Dis 2023: 9551163. doi:10.1155/2023/9551163

2. Ahmed Z, Renart EG, Zeeshan S. 2021. Genomics pipelines to investigate susceptibility in whole genome and exome sequenced data for variant discovery, annotation, prediction and genotyping. PeerJ 9: e11724. doi:10.7717/peerj.11724

3. Ahsan MM, Luna SA, Siddique Z. 2022. Machine-learning-based disease diagnosis: A comprehensive review. Healthcare 10: 541. doi:10.3390/healthcare10030541

4. Alzubi R, Ramzan N, Alzoubi H, Amira A. 2017. A hybrid feature selection method for complex diseases SNPs. IEEE Access 6: 1292–1301. doi:10.1109/ACCESS.2017.2778268

5. Anastassopoulou C, Davaris N, Ferous S, Siafakas N, Boufidou F, Anagnostopoulos K, Tsakris A. 2024. The Molecular Basis of Olfactory Dysfunction in COVID-19 and Long COVID. Lifestyle Genom 17: 42–56. doi:10.1159/000539292

6. Arslan, E, Schulz, J, Rai K. 2021. Machine learning in epigenomics: Insights into cancer biology and medicine. Biochim Biophys Acta Rev Cancer 1876: 188588. doi:10.1016/j.bbcan.2021.188588

7. Augusto DG, Murdolo LD, Chatzileontiadou DS, Sabatino Jr JJ, Yusufali T, Peyser ND, Butcher X, Kizer K, Guthrie K, Murray VW, et al. 2023. A common allele of HLA is associated with asymptomatic SARS-CoV-2 infection. Nature 620: 128–136. doi:10.1038/s41586-023-06331-x

8. Behravan H, Hartikainen JM, Tengström M, Pylkäs K, Winqvist R, Kosma VM, Mannermaa A. 2018. Machine learning identifies interacting genetic variants contributing to breast cancer risk: A case study in Finnish cases and controls. Sci Rep 8: 13149. doi:10.1038/s41598-018-31573-5

9. Berkowicz DA, Trombley PQ, Shepherd GM. 1994. Evidence for glutamate as the olfactory receptor cell neurotransmitter. J Neurophysio 71: 2557–2561. doi:10.1152/jn.1994.71.6.2557

10. Bicker F, Vasic V, Horta G, Ortega F, Nolte H, Kavyanifar A, Keller S, Stankovic ND, Harter PN, Benedito R, et al. Neurovascular EGFL7 regulates adult neurogenesis in the subventricular zone and thereby affects olfactory perception. Nat Commun 8: 15922. doi:10.1038/ncomms15922

11. Butowt R, Bilinska K, von Bartheld CS. 2023. Olfactory dysfunction in COVID-19: new insights into the underlying mechanisms. Trends Neurosci 46: 75–90. doi:10.1016/j.tins.2022.11.003

12. Butowt R, von Bartheld CS. 2021. Anosmia in COVID-19: underlying mechanisms and assessment of an olfactory route to brain infection. Neuroscientist 27: 582–603. doi:10.1177/1073858420956905

13. Carithers LJ, Moore HM. 2015. The genotype-tissue expression (GTEx) project. Biopreserv Biobank 13: 307. doi:10.1089/bio.2015.29031.hmm

14. Cazzolla, AP, Lovero R, Lo Muzio L, Testa NF, Schirinzi A, Palmieri G, Pozzessere P, Procacci V, Di Comite M, Ciavarella D, et al. 2020. Taste and smell disorders in COVID-19 patients: role of interleukin-6. ACS Chem Neurosci 11: 2774–2781. doi:10.1021/acschemneuro.0c00447

15. Centers for Disease Control and Prevention. 2024, May 23. COVID Data Tracker. Atlanta, GA: U.S. Department of Health and Human Services, CDC. https://covid.cdc.gov/covid-data-tracker

16. Chang CC, Chow CC, Tellier LC, Vattikuti S, Purcell SM, Lee JJ. 2015. Second-generation PLINK: rising to the challenge of larger and richer datasets. Gigascience 4: s13742–015. doi:10.1186/s13742-015-0047-8

17. Christov CP, Gardiner TJ, Szüts D, Krude T. 2006. Functional requirement of noncoding Y RNAs for human chromosomal DNA replication. Mol Cell Biol 26: 6993–7004. doi:10.1128/MCB.01060-06

18. Chung J, Vig V, Sun X, Han X, O’Connor GT, Chen X, DeAngelis MM, Farrer LA, Subramanian ML. 2022. Genome-wide pleiotropy study identifies association of PDGFB with age-related macular degeneration and COVID-19 infection outcomes. J Clin Med 12: 109. doi:10.3390/jcm12010109

19. Corey EA, Ukhanov K, Bobkov YV, McIntyre JC, Martens JR, Ache BW. 2021. Inhibitory signaling in mammalian olfactory transduction potentially mediated by Gαo. Mol Cell Neurosci 110: 103585. doi:10.1016/j.mcn.2020.103585

20. COVID-19 Host Genetics Initiative. 2021. Mapping the human genetic architecture of COVID-19. Nature 600: 472–477. doi:10.1038/s41586-021-03767-x

21. Danecek P, Bonfield JK, Liddle J, Marshall J, Ohan V, Pollard MO, Whitwham A, Keane T, McCarthy SA, Davies RM, et al. 2021. Twelve years of SAMtools and BCFtools. Gigascience 10: giab008. doi:10.1093/gigascience/giab008

22. Darapaneni V, Jaldani A. 2021. Membrane protein of SARS-CoV-2 plays a pivotal role in the availability of active testosterone through its interaction with AKR1C2 enzyme leading to the upregulation of TMPRSS2 protease expression. Microbiol Indep Res J 8: 55–57. doi:10.18527/2500-2236-2021-8-1-38-40

23. Davis GE. 2010. Vascular balancing act: EGFL7 and Notch. Blood 116: 5791–5793. doi:10.1182/blood-2010-11-314500

24. Diepeveen J, Moerdijk-Poortvliet TC, van der Leij FR. 2022. Molecular insights into human taste perception and umami tastants: A review. J Food Sci 87: 1449–1465. doi:10.1111/1750-3841.16101

25. Doege H, Bocianski A, Scheepers A, Axer H, Eckel J, Joost HG, Schürmann A. 2001. Characterization of human glucose transporter (GLUT) 11 (encoded by SLC2A11), a novel sugar-transport facilitator specifically expressed in heart and skeletal muscle. Biochem J 359: 443–449. doi:10.1042/bj3590443

26. Ewels P, Magnusson M, Lundin S, Käller M. 2016. MultiQC: summarize analysis results for multiple tools and samples in a single report. Bioinformatics 32: 3047–3048. doi:10.1093/bioinformatics/btw354

27. Finlay JB, Brann DH, Abi Hachem R, Jang DW, Oliva DW, Ko T, Gupta R, Wellford SA, Moseman EA, Jang SS, et al. 2022. Persistent post–COVID-19 smell loss is associated with immune cell infiltration and altered gene expression in olfactory epithelium. Sci Transl Med 14: eadd0484. doi:10.1126/scitranslmed.add0484

28. Fisher A, Rudin C, Dominici F. 2019. All models are wrong, but many are useful: Learning a variable’s importance by studying an entire class of prediction models simultaneously. J Mach Learn Res 20: 1–81.

29. Frankish A, Diekhans M, Jungreis I, Lagarde J, Loveland JE, Mudge JM, Sisu C, Wright JC, Armstrong J, Barnes I, et al. 2021. GENCODE 2021. Nucleic Acids Res 49: D916–D923. doi:10.1093/nar/gkaa1087

30. Genovese G, Rockweiler NB, Gorman BR, Bigdeli TB, Pato MT, Pato CN, Ichihara K, McCarroll SA. 2024. BCFtools/liftover: an accurate and comprehensive tool to convert genetic variants across genome assemblies. Bioinformatics 40: btae038. doi:10.1093/bioinformatics/btae038

31. Goldstein BA, Polley EC, Briggs FB. 2011. Random forests for genetic association studies. Stat Appl Genet Mol Biol 10. doi:10.2202/1544-6115.1691

32. Guo P, Hu B, Gu W, Xu L, Wang D, Huang HJ, Cavenee WK, Cheng SY. 2003. Platelet-derived growth factor-B enhances glioma angiogenesis by stimulating vascular endothelial growth factor expression in tumor endothelia and by promoting pericyte recruitment. Am J Pathol 162: 1083–1093. doi:10.1016/S0002-9440(10)63905-3

33. Henarejos-Castillo I, Aleman A, Martinez-Montoro B, Gracia-Aznárez FJ, Sebastian-Leon P, Romeu M, Remohi J, Patiño-Garcia A, Royo P, Alkorta-Aranburu G, et al. 2021. Machine learning-based approach highlights the use of a genomic variant profile for precision medicine in ovarian failure. J Pers Med 11: 609. doi:10.3390/jpm11070609

34. Ho DSW, Schierding W, Wake M, Saffery R, O’Sullivan J. 2019. Machine learning SNP based prediction for precision medicine. Front Genet 10: 431037. doi:10.3389/fgene.2019.00267

35. Iosef C, Martin CM, Slessarev M, Gillio-Meina C, Cepinskas G, Han VK, Fraser DD. 2023. COVID-19 plasma proteome reveals novel temporal and cell-specific signatures for disease severity and high-precision disease management. J Cell Mol Med 27: 141–157. doi:10.1111/jcmm.17622

36. Jin H, Ahn J, Park Y, Sim J, Park HS, Ryu CS, Kim NK, Kwack K. 2020. Identification of potential causal variants for premature ovarian failure by whole exome sequencing. BMC Med Genet 13: 1–8. doi:10.1186/s12920-020-00813-x

37. Jin M, Yu B, Zhang W, Zhang W, Xiao Z, Mao Z, Lai Y, Lin D, Ma Q, Pan E, et al. 2016. Toll-like receptor 2-mediated MAPKs and NF-κsB activation requires the GNAO1-dependent pathway in human mast cells. Integr Biol 8: 968–975. doi:10.1039/c6ib00097e

38. Karolchik D, Hinrichs AS, Furey TS, Roskin KM, Sugnet CW, Haussler D, Kent WJ. 2004. The UCSC Table Browser data retrieval tool. Nucleic Acids Res 32: D493–D496. doi:10.1093/nar/gkh103

39. Komuro A, Masuda Y, Kobayashi K, Babbitt R, Gunel M, Flavell RA, Marchesi VT. 2004. The AHNAKs are a class of giant propeller-like proteins that associate with calcium channel proteins of cardiomyocytes and other cells. Proc Natl Acad Sci USA 101: 4053–4058. doi:10.1073/pnas.030861910

40. Krämer A, Green J, Pollard Jr J, Tugendreich S. 2014. Causal analysis approaches in ingenuity pathway analysis. Bioinformatics 30: 523–530. doi:10.1093/bioinformatics/btt703

41. Krishnakumar HN, Momtaz DA, Sherwani A, Mhapankar A, Gonuguntla RK, Maleki A, Abbas A, Ghali AN, Al Afif A. 2023. Pathogenesis and progression of anosmia and dysgeusia during the COVID-19 pandemic. Eur Arch Otorhinolaryngol 280: 505–509. doi:10.1007/s00405-022-07689-w

42. Kristiansen H, Gad HH, Eskildsen-Larsen S, Despres P, Hartmann R. 2011. The oligoadenylate synthetase family: an ancient protein family with multiple antiviral activities. J Interferon Cytokine Res 31: 41–47. doi:10.1089/jir.2010.010

43. Kuhn M, Wickham H, Hvitfeldt E. 2024. recipes: Preprocessing and Feature Engineering Steps for Modeling. R package version 1.0.10, https://recipes.tidymodels.org/, https://github.com/tidymodels/recipes

44. Kumar AA, Lee SW, Lock C, Keong NC. 2021. Geographical variations in host predisposition to COVID-19 related anosmia, ageusia, and neurological syndromes. Front Med 8: 661359. doi:10.3389/fmed.2021.661359

45. Kursa MB. 2021. Praznik: High performance information-based feature selection. SoftwareX, 16: 100819. doi:10.1016/j.softx.2021.100819.

46. Lello L, Raben TG, Yong SY, Tellier LC, Hsu SD. 2019. Genomic prediction of 16 complex disease risks including heart attack, diabetes, breast and prostate cancer. Sci Rep 9: 15286. doi: 10.1038/s41598-019-51258-x

47. Liu L, Meng Q, Weng C, Lu Q, Wang T, Wen Y. 2022. Explainable deep transfer learning model for disease risk prediction using high-dimensional genomic data. PLoS Comput Biol 18: e1010328. doi:10.1371/journal.pcbi.1010328

48. Magnusson M. 2016. VCF Parser. GitHub repository. https://github.com/moonso/vcf_parser

49. Mahmoud MM, Abuohashish HM, Khairy DA, Bugshan AS, Khan AM, Moothedath MM. 2021. Pathogenesis of dysgeusia in COVID-19 patients: a scoping review. Eur Rev Med Pharmacol Sci 25: 1114–1134. doi:10.26355/eurrev_202101_24683

50. Matuozzo D, Talouarn E, Marchal A, Zhang P, Manry J, Seeleuthner Y, Zhang Y, Bolze A, Chaldebas M, Milisavljevic B, et al. 2023. Rare predicted loss-of-function variants of type I IFN immunity genes are associated with life-threatening COVID-19. Genome Med 15: 22. doi:10.1186/s13073-023-01173-8

51. McKibbin W, Fernando R. 2023. The global economic impacts of the COVID-19 pandemic. Econ Model 129: 106551. doi:10.1016/j.econmod.2023.106551

52. McLaren W, Gil L, Hunt SE, Riat HS, Ritchie GR, Thormann A, Flicek P, Cunningham F. 2016. The ensembl variant effect predictor. Genome Biol 17: 1–14. doi:10.1186/s13059-016-0974-4

53. Mick E, Kamm J, Pisco AO, Ratnasiri K, Babik JM, Castañeda G, DeRisi JL, Detweiler AM, Hao SL, Kangelaris KN, et al. 2020. Upper airway gene expression reveals suppressed immune responses to SARS-CoV-2 compared with other respiratory viruses. Nat Commun 11: 5854. doi:10.1038/s41467-020-19587-y

54. Miwa T, Mori E, Sekine R, Kimura Y, Kobayashi M, Shiga H, Tsuzuki K, Suzuki M, Kondo K, Suzaki I, et al. 2023. Olfactory and taste dysfunctions caused by COVID-19: a nationwide study. Rhinology 61: 552–560. doi:10.4193/Rhin23.034

55. Mohar NP, Cox EM, Adelizzi E, Moore SA, Mathews KD, Darbro BW, Wallrath LL. 2024. The Influence of a Genetic Variant in CCDC78 on LMNA-Associated Skeletal Muscle Disease. Int J Mol Sci 25: 4930. doi:10.3390/ijms25094930

56. Mollica V, Rizzo A, Massari F. 2020. The pivotal role of TMPRSS2 in coronavirus disease 2019 and prostate cancer. Future Oncol 16: 2029–2033. doi:10.2217/fon-2020-0571

57. Musolf AM, Holzinger ER, Malley JD, Bailey-Wilson JE. 2022. What makes a good prediction? Feature importance and beginning to open the black box of machine learning in genetics. Hum Genet 141: 1515–1528. doi:10.1007/s00439-021-02402-z

58. Nicholls HL, John CR, Watson DS, Munroe PB, Barnes MR, Cabrera CP. 2020. Reaching the end-game for GWAS: machine learning approaches for the prioritization of complex disease loci. Front Genet 11: 521712. doi:10.3389/fgene.2020.00350

59. Nicola M, Alsafi Z, Sohrabi C, Kerwan A, Al-Jabir A, Iosifidis C, Agha M, Agha R. 2020. The socio-economic implications of the coronavirus pandemic (COVID-19): A review. Int Surg J 78: 185–193. doi:10.1016/j.ijsu.2020.04.018

60. Okamoto H, Ichikawa N. 2021. The pivotal role of the angiotensin-II–NF-κB axis in the development of COVID-19 pathophysiology. Hypertens Res 44: 126–128. doi:10.1038/s41440-020-00560-7

61. Olliff NS, Hunt MA, Paudel SS, Nguyen KN, Delcher HA, DeMeis JD, Roberts JT, Fouty BW, Audia JP, Kim JH, et al. 2023. Human YRNA 4 (HY4) plasma levels are a prognostic indicator of SARS-CoV-2 infection clinical severity. MicroPubl Biol 2023. doi:10.17912/micropub.biology.000925

62. Orrù V, Steri M, Sidore C, Marongiu M, Serra V, Olla S, Sole G, Lai S, Dei M, Mulas A, et al. 2020. Complex genetic signatures in immune cells underlie autoimmunity and inform therapy. Nat Genet 52: 1036–1045. doi:10.1038/s41588-020-0684-4

63. Penning TM, Burczynski ME, Jez JM, Hung CF, Lin HK, Ma H, Moore M, Palackal N, Ratnam K. 2000. Human 3α-hydroxysteroid dehydrogenase isoforms (AKR1C1–AKR1C4) of the aldo-keto reductase superfamily: functional plasticity and tissue distribution reveals roles in the inactivation and formation of male and female sex hormones. Biochem J 351: 67–77. doi:10.1042/bj3510067

64. Pinnaro CT, Beck CB, Major HJ, Darbro BW. 2023. CRELD1 variants are associated with bicuspid aortic valve in Turner syndrome. Hum Genet 142: 523–530. doi:10.1007/s00439-023-02538-0

65. Pinnaro CT, Henry T, Major HJ, Parida M, DesJardin LE, Manak JR, Darbro BW. 2020. Candidate modifier genes for immune function in 22q11. 2 deletion syndrome. Mol Genet Genomic Med 8: e1057. doi:10.1002/mgg3.1057

66. Python Software Foundation. 2023. Python Language Reference, version 3.10.12. http://www.python.org/

67. R Core Team. 2022. R: A Language and Environment for Statistical Computing. R Foundation for Statistical Computing: Vienna, Austria. https://www.R-project.org/

68. Reel, PS, Reel S, Pearson E, Trucco E, Jefferson E. 2021. Using machine learning approaches for multi-omics data analysis: A review. Biotechnol Adv 49: 107739. doi:10.1016/j.biotechadv.2021.107739

69. Rentzsch P, Witten D, Cooper GM, Shendure J, Kircher M. 2019. CADD: predicting the deleteriousness of variants throughout the human genome. Nucleic Acids Res 47: D886–D894. doi:10.1093/nar/gky1016

70. Rižner TL, Penning TM. 2014. Role of aldo–keto reductase family 1 (AKR1) enzymes in human steroid metabolism. Steroids 79: 49–63. doi:10.1016/j.steroids.2013.10.012

71. Romagnoni A, Jégou S, Van Steen K, Wainrib G, Hugot JP. 2019. Comparative performances of machine learning methods for classifying Crohn Disease patients using genome-wide genotyping data. Sci Rep 9: 10351. doi:10.1038/s41598-019-46649-z

72. Roper SD. 2013. Taste buds as peripheral chemosensory processors. Semin Cell Dev Biol 24: 71–79. doi:10.1016/j.semcdb.2012.12.002

73. Samavarchi-Tehrani P, Abdouni H, Knight JD, Astori A, Samson R, Lin ZY, Kim DK, Knapp JJ, St-Germain J, Go CD, et al. 2020. A SARS-CoV-2–host proximity interactome. *BioRxiv* doi:10.1101/2020.09.03.282103

74. Sanchez A, Kuras M, Murillo JR, Pla I, Pawlowski K, Szasz AM, Gil J, Nogueira FC, Perez-Riverol Y, Eriksson J, et al. 2020. Novel functional proteins coded by the human genome discovered in metastases of melanoma patients. Cell Biol Toxicol 36: 261–272. doi:10.1007/s10565-019-09494-4

75. Sardaar S, Qi B, Dionne-Laporte A, Rouleau GA, Rabbany R, Trakadis YJ. 2020. Machine learning analysis of exome trios to contrast the genomic architecture of autism and schizophrenia. Bmc Psychiatry 20: 1–11. doi:10.1186/s12888-020-02503-5

76. Severe Covid-19 GWAS Group. 2020. Genomewide association study of severe Covid-19 with respiratory failure. N Engl J Med 383: 1522–1534. doi:10.1056/NEJMoa2020283

77. Shelton, JF, Shastri AJ, Fletez-Brant K, Aslibekyan S, Auton A. 2022. The UGT2A1/UGT2A2 locus is associated with COVID-19-related loss of smell or taste. Nat Genet 54: 121–124. doi:10.1038/s41588-021-00986-w

78. Smith BJ. 2024. MachineShop: Machine Learning Models and Tools. R Package Version 3.7.0. https://cran.r-project.org/package=MachineShop

79. Sun H, Lan X, Ma L, Zhou J. 2022. Revealing modifier variations characterizations for elucidating the genetic basis of human phenotypic variations. Hum Genet 141: 1223–1233. doi:10.1007/s00439-021-02362-4

80. Trakadis YJ, Sardaar S, Chen A, Fulginiti V, Krishnan A. 2019. Machine learning in schizophrenia genomics, a case-control study using 5,090 exomes. Am J Med Genet B Neuropsychiatr Genet 180: 103–112. doi:10.1002/ajmg.b.32638

81. Usuba R, Pauty J, Soncin F, Matsunaga YT. 2019. EGFL7 regulates sprouting angiogenesis and endothelial integrity in a human blood vessel model. Biomaterials 197: 305–316. doi:10.1016/j.biomaterials.2019.01.022

82. Vabalas A, Gowen E, Poliakoff E, Casson AJ. 2019. Machine learning algorithm validation with a limited sample size. PloS one 14: e0224365. doi:10.1371/journal.pone.0224365

83. Vadapalli S, Abdelhalim H, Zeeshan S, Ahmed Z. 2022. Artificial intelligence and machine learning approaches using gene expression and variant data for personalized medicine. Brief Bioinform 23: bbac191. doi:10.1093/bib/bbac191

84. Van der Auwera GA, O’Connor BD. 2020. *Genomics in the cloud: using Docker, GATK, and WDL in Terra*. O’Reilly Media, Sebastopol, California.

85. Weiner DJ, Nadig A, Jagadeesh KA, Dey KK, Neale BM, Robinson EB, Karczewski KJ, O’Connor LJ. 2023. Polygenic architecture of rare coding variation across 394,783 exomes. Nature 614: 492–499. doi: 10.1038/s41586-022-05684-z

86. World Health Organization. 2024, May 23. WHO Coronavirus (COVID-19) dashboard > Cases [Dashboard]. https://data.who.int/dashboards/covid19/cases

87. Wynne C, Lazzari E, Smith S, McCarthy EM, Ní Gabhann J, Kallal LE, Higgs R, Cryan SA, Biron CA, Jefferies CA. 2014. TRIM68 negatively regulates IFN-β production by degrading TRK fused gene, a novel driver of IFN-β downstream of anti-viral detection systems. PloS one 9: e101503. doi:10.1371/journal.pone.0101503

88. Yang QZ, Porter BE, Axeen ET. 2023. GNAO1-related neurodevelopmental disorder: Literature review and caregiver survey. Epilepsy Behav Rep 21: 100582. doi:10.1016/j.ebr.2022.100582

89. Yuki K, Fujiogi M, Koutsogiannaki S. 2020. COVID-19 pathophysiology: A review. J Clin Immunol 215: 108427. doi:10.1016/j.clim.2020.108427

90. Zguro K, Fallerini C, Fava F, Furini S, Renieri A. 2022. Host genetic basis of COVID-19: from methodologies to genes. Eur J Hum Genet 30: 899–907. doi:10.1038/s41431-022-01121-x

91. Zhang J, Mai S, Chen HM, Kang K, Li XC, Chen SH, Pan PY. 2017. Leukocyte immunoglobulin-like receptors in human diseases: an overview of their distribution, function, and potential application for immunotherapies. J Leukoc Biol 102: 351–360. doi:10.1189/jlb.5MR1216-534R

92. Zhang T, Kutler D, Scognamiglio T, Gudas LJ, Tang XH. 2023. Transcriptomic analysis predicts the risk of progression of premalignant lesions in human tongue. Discov Oncol 14: 24. doi:10.1007/s12672-023-00629-y

