## Supplemental Materials and Figures for "Genome-wide Machine Learning Analysis of Anosmia and Ageusia with COVID-19"

[1]Interdisciplinary Graduate Program in Genetics, University of Iowa, Iowa City, IA 52242, USA;
[2]Department of Biomedical Engineering, University of Iowa, Iowa City, IA 52242, USA; (L.P.); (T.B.)
[3]Stead Family Department of Pediatrics, University of Iowa, Iowa City, IA 52242, USA; (E.P.); (M.M.); (H.E.); (B.D.)
[4]Department of Biostatistics, University of Iowa, Iowa City, IA 52242, USA; (B.J.S)
[5]Center for Bioinformatics and Computational Biology, University of Iowa, Iowa City, IA 52242, USA;
[6]Department of Electrical and Computer Engineering, University of Iowa, Iowa City, IA 52242, USA; (T.C.)

### **SUPPLEMENTARY MATERIALS AND METHODS S1**

Description of additional feature selection pipelines examined.

Best filtering practices were tested on the UI cohort. The order of the filtering methods in the pipeline was determined by initial preliminary results considering the number of features passing the filters and the computation time. DT-VI has a higher computational load than CMI and LR filtering leading to CMI and LR methods to be performed first. Our initial pipeline tested was CMI-5, LR (chi-squared, p-value threshold < 0.1), and DT-VI_1000_1 filtering (majority class accuracy threshold) to confirm this strategy could yield informative machine learning results. Multiple thresholds for CMI and LR methods were tested individually. For CMI, the number of features selected for each intermediate dataset were 5, 10, 20, 100, and 500 features. When applicable, input of the complete dataset was used for CMI filtering selecting 500 features (CMI-Full). For LR, p-values thresholds of 0.1, 0.05, 0.01, 0.005, 0.001, 0.0005, 0.0001, 0.00005, and 0.00001 were tested for chi-squared test and F-test statistics. LR as the initial filtering method was also tested. Testing was performed with model-based variable importance filtering as the initial filter similar to DT-VI_1000_1 with different models. Models tested were DT, RF, Lasso, NB, SVM-L, and XGBTree model with a training set majority class accuracy threshold and variable importance score set to either > 0 or > 75. The RF and Lasso filters were also tested with accuracy thresholds of 60% and variable importance score > 50.

Datasets from the best performing individual filter results and a combination of CMI and LR filtering results were used as input to DT-VI_1000_1 and DT-VI_1000_10 filtering and tested for predictive performance with ML models. DT-VI_X_10 was tested in multiple ways. We assessed the number of features sampled from the initial training set including X = 50, 100, 250, 500, and 1000. We expanded on the DT-VI_50_10 approach by testing accuracy thresholds in two ways, the first by maintaining the same input dataset and testing the accuracy thresholds of majority class (~53%), 60%, 65%, 70%, 75%, and 80%. The second test examined increasing accuracy thresholds in the same manner as the first but using the prior accuracy threshold filtered output dataset as the input dataset to the next test. We expanded on the DT-VI_1000_10 filtering approach by assessing different 10-fold CV estimated performance metrics to select features, with and without allele features converted to factors. The metrics and thresholds assessed were accuracy (thresholds described previously), brier score < 0.25, 0.3, AUROC curve > 0.55, 0.65, sensitivity > 0.5, 0.75, and specificity > 0.5, 0.75. Output datasets from sensitivity and specificity tests were combined to assess the viability of combination effects of features on performance metrics.

The Gene feature analysis tested multiple filtering strategies similar to the variant feature analysis. We used the initial variant feature dataset with NC80 and ZV filtering as the input dataset to the gene feature analysis pipeline. Preliminary analysis examined the performance of transforming the initial training dataset into gene features without any filtering and demonstrated the necessity for filtering before the gene feature transformation. We examined multiple filtering strategies before transformation with both corrections and with no correction including single filters with CMI-20 and LR (p-value threshold < 0.1, 0.05, 0.005) and two filters with all applicable permutations of CMI-20 and LR (p-value threshold < 0.1, 0.05, 0.005) along with DT-VI_1000_1 for the second filter. We expanded the single filter analysis by including a second filter after the gene feature transformation with all permutations of CMI-20 and LR (p-value threshold < 0.1, 0.05, 0.005) for the first filter and CMI-20, CMI-Full, LR (p-value threshold < 0.1, 0.05, 0.01, 0.005, 0.001), DT-VI_1000_1, and DT-VI_50_1 for the gene feature (second) filter. All filtering approaches were assessed by predictive performance of models trained to the data as previously described. A complete list of feature selection pipelines tested is included in Supplementary Table S9.

### SUPPLEMENTARY TABLES

Refer to Supplemental_Tables.xlsx for supplemental tables and legends.

**Supplementary Table S1.** Summary statistics for the University of Iowa (UI) cohort.

**Supplementary Table S2.** Individual Covariate Association Test with Response Variables.

**Supplementary Table S3.** UI cohort loss of smell only phenotype coding and noncoding dataset initial pipeline results. CMI-5, LR, DT-VI_1000_1.

**Supplementary Table S4.** UI cohort loss of smell only phenotype coding regions only dataset initial pipeline results. CMI-5, LR, DT-VI_1000_1.

**Supplementary Table S5.** Lists of top datasets and variables from the variant and gene analyses of the UI and AoU cohorts.

**Supplementary Table S6.** UI cohort loss of smell only phenotype top variant analysis results. CMI-20, DT-VI_1000_10.

**Supplementary Table S7.** Permutation-based variable importance (PVI) all features for top performing dataset (CMI-20, DT-VI_1000_10) and all models for the UI cohort loss of smell only variant analysis. Permutations per feature were performed 25 times and performance is assessed based on mean brier score.

**Supplementary Table S8.** UI cohort loss of smell only phenotype top gene analysis results. LR (p-value threshold = 0.05), gene feature transformation with the sample allele frequency correction, CMI-20.

**Supplementary Table S9.** Permutation-based variable importance (PVI) all features for top performing dataset (LR (p-value threshold = 0.05), gene feature transformation with the sample allele frequency correction, CMI-20) and all models for the UI cohort loss of smell only gene analysis. Permutations per feature were performed 25 times and performance is assessed based on mean brier score.

**Supplementary Table S10.** Complete list of feature selection pipelines examined on the UI anosmia only dataset.

**Supplementary Table S11.** Complete 10-fold cross validation training performance and held-out test results for all variant and gene feature selection strategies and pipelines for the UI loss of smell only phenotype.

**Supplementary Table S12.** UI cohort loss of taste only phenotype top variant analysis results. CMI-5.

**Supplementary Table S13.** Permutation-based variable importance (PVI) all features for top performing dataset (CMI-5) and all models for the UI cohort loss of taste only variant analysis. Permutations per feature were performed 25 times and performance is assessed based on mean brier score.

**Supplementary Table S14.** UI cohort loss of taste only phenotype top gene analysis results. LR (p-value threshold = 0.05, chi-squared), gene variable transformation with the directional correction, CMI-20.

**Supplementary Table S15.** Permutation-based variable importance (PVI) all features for top performing dataset (LR (p-value threshold = 0.05), gene variable transformation with the directional correction, CMI-20) and all models for the UI cohort loss of taste only gene analysis. Permutations per feature were performed 25 times and performance is assessed based on mean brier score.

**Supplementary Table S16.** Complete 10-fold cross validation training performance and held-out test results for all variant and gene feature selection strategies and pipelines for the UI loss of taste only phenotype.

**Supplementary Table S17.** UI cohort loss of smell and/or taste phenotype top variant analysis results. CMI-5, LR (p-value threshold = 0.1, F test), DT-VI_1000_1.

**Supplementary Table S18.** Permutation-based variable importance (PVI) all features for top performing dataset (CMI-5, LR (p-value threshold = 0.1, F test), DT-VI_1000_1) and all models for the UI cohort loss of smell and/or taste variant analysis. Permutations per feature were performed 25 times and performance is assessed based on mean brier score.

**Supplementary Table S19.** UI cohort loss of smell and/or taste phenotype top gene analysis results. LR (p-value threshold = 0.05, chi-squared), gene variable transformation with the sample frequency correction, CMI-20.

**Supplementary Table S20.** Permutation-based variable importance (PVI) all features for top performing dataset (LR (p-value threshold = 0.05, chi-squared), gene variable transformation with the sample frequency correction, CMI-20) and all models for the UI cohort loss of smell and/or taste gene analysis. Permutations per feature were performed 25 times and performance is assessed based on mean brier score.

**Supplementary Table S21.** Complete 10-fold cross validation training performance and held-out test results for all variant and gene feature selection strategies and pipelines for the UI loss of smell and/or taste phenotype.

**Supplementary Table S22.** Intersection of features, genomic positions, and genes of the top performing datasets from the three phenotype analyses from the UI cohort and AoU cohort.

**Supplementary Table S23.** Summary statistics for the All of Us (AoU) cohort.

**Supplementary Table S24.** Model performance of top datasets and models from the UI cohort variant and gene analysis tested on the AoU cohort.

**Supplementary Table S25.** Permutation test between group identity-by-state differences with plotting PC1 and PC2 from principal component analysis.

**Supplementary Table S26.** Complete 10-fold cross validation training performance and held-out test results for all variant and gene feature selection strategies and pipelines for the AoU cohort.

**Supplementary Table S27.** Permutation-based variable importance (PVI) all features for top performing dataset (CMI-5, LR (p-value threshold = 0.1, chi-squared), DT-VI_1000_1 + CMI-5, LR (p-value threshold = 0.1, F test), DT-VI_1000_1 with Covariates) and all models for the AoU variant analysis. Permutations per feature were performed 25 times and performance is assessed based on mean brier score.

**Supplementary Table S28.** Permutation-based variable importance (PVI) all features for top performing dataset (LR (p-value threshold = 0.05, chi-squared), gene variable transformation with no correction, CMI-20 with Covariates) and all models for the AoU gene analysis. Permutations per feature were performed 25 times and performance is assessed based on mean brier score.

**Supplementary Table S29.** Model performance of the top dataset and models from the AoU cohort variant analysis tested on the UI cohort.

**Supplementary Table S30.** Association test of previously implicated variants for loss of smell and/or taste with COVID-19 in the UI and AoU cohorts.

**Supplementary Table S31.** IPA individual dataset Core Canonical Pathway Analaysis for top datasets from the UI and AoU cohorts.

**Supplementary Table S32.** IPA Comparison Analysis of Core Canonical Pathway for top datasets from the UI and AoU cohorts.

**Supplementary Table S33.** IPA Comparison Analysis of Core Upstream Regulators for top datasets from the UI and AoU cohorts.

**Supplementary Table S34.** Models and parameter values for hyperparameter tuning. Parameters were tuned with a grid search and 10-fold CV with training sets.

### SUPPLEMENTARY FIGURES

**A**

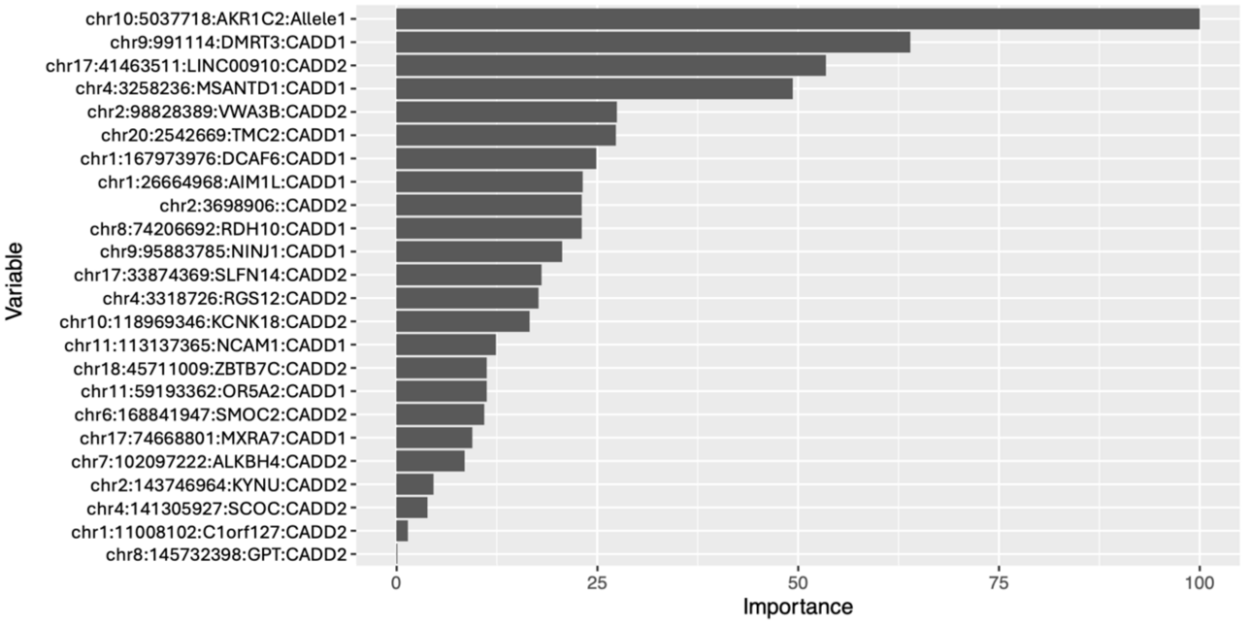

**B**

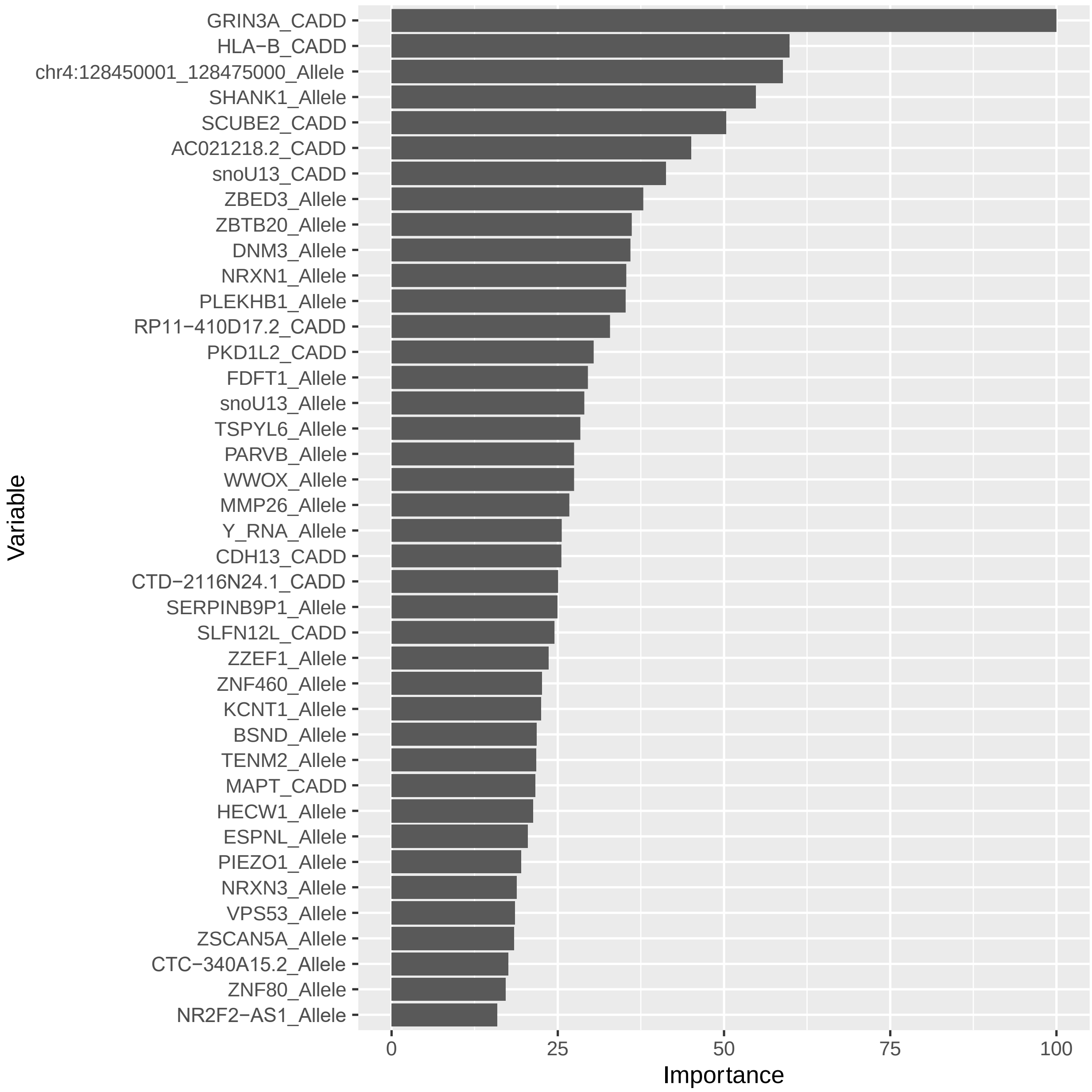
GRIN3A_CADD
HLA−B_CADD
chr4:128450001_128475000_Allele
SHANK1_Allele
SCUBE2_CADD
AC021218.2_CADD
snoU13_CADD
ZBED3_Allele
ZBTB20_Allele
DNM3_Allele
NRXN1_Allele
PLEKHB1_Allele
RP11−410D17.2_CADD
PKD1L2_CADD
FDFT1_Allele
snoU13_Allele
TSPYL6_Allele
PARVB_Allele
WWOX_Allele
MMP26_Allele
Y_RNA_Allele
CDH13_CADD
CTD−2116N24.1_CADD
SERPINB9P1_Allele
SLFN12L_CADD
ZZEF1_Allele
ZNF460_Allele
KCNT1_Allele
BSND_Allele
TENM2_Allele
MAPT_CADD
HECW1_Allele
ESPNL_Allele
PIEZO1_Allele
NRXN3_Allele
VPS53_Allele
ZSCAN5A_Allele
CTC−340A15.2_Allele
ZNF80_Allele
NR2F2−AS1_Allele
Variable
0
25
50
75
100
Importance

C

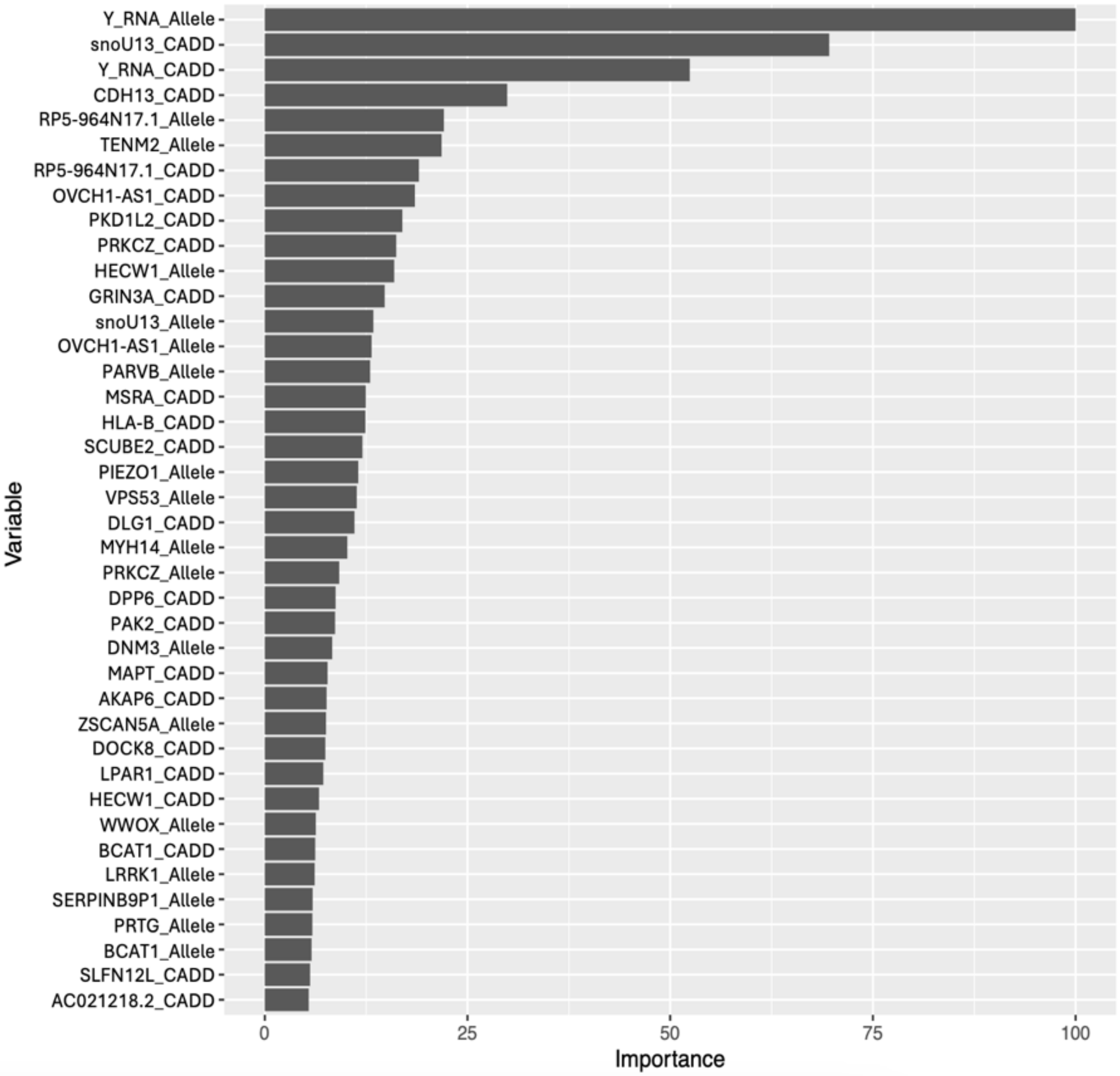

**Supplementary Figure S1**. Permutation-based variable importance top 40 features for top performing dataset and models for the UI cohort loss of taste analysis. All features included in the model are assessed. (*A*) Variable importance for the decision tree (DT) model, top performing model for the variant analysis. (*B*) Variable importance for the support vector machine with linear kernel function (SVM-L) model, top performing model for the gene analysis according to the accuracy and AUROC curve metrics. (*C*) Variable importance for the RF model, top performing model for the gene analysis according to brier score.

**A**

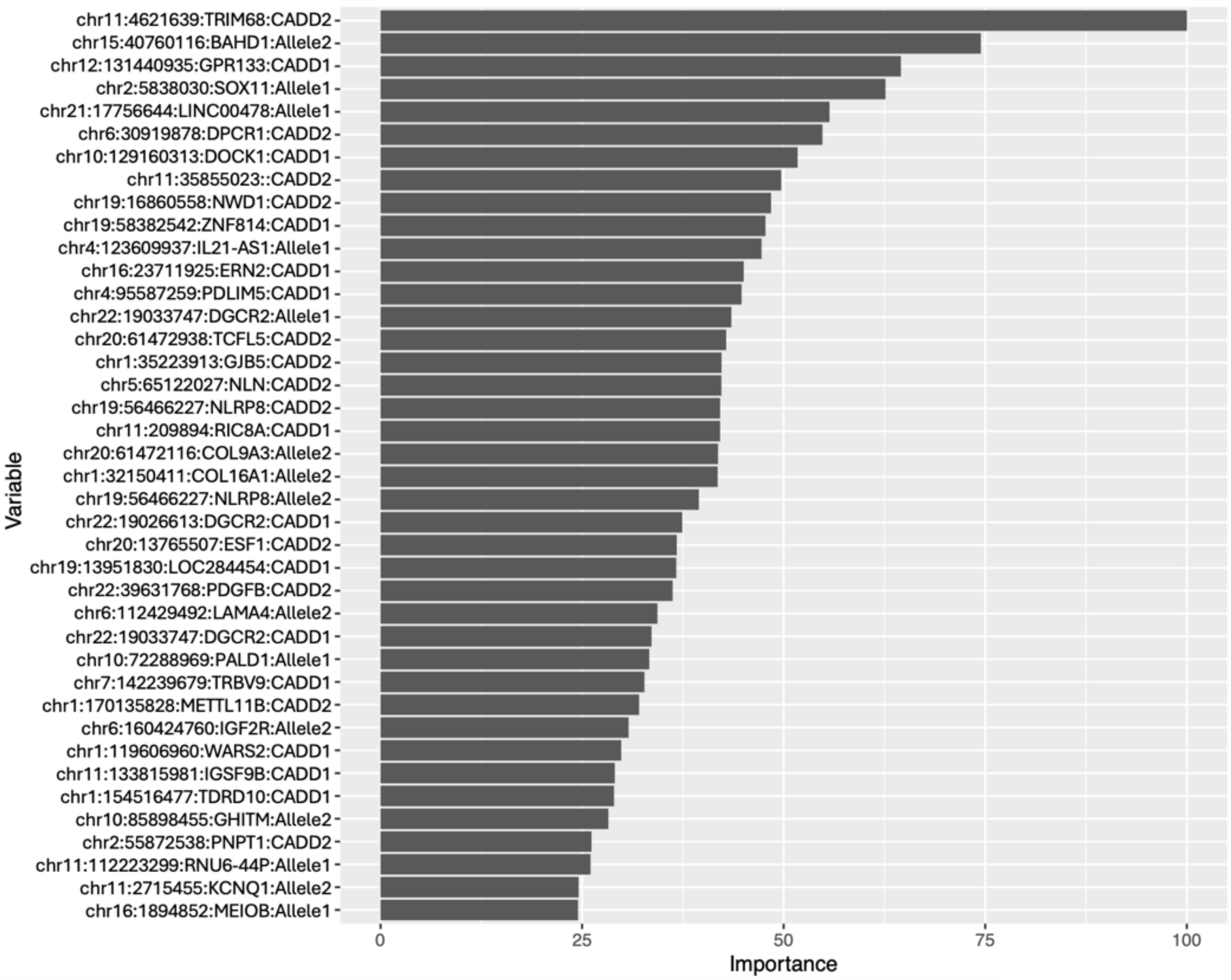
chr11:4621639:TRIM68:CADD2
chr15:40760116:BAHD1:Allele2
chr12:131440935:GPR133:CADD1
chr2:5838030:SOX11:Allele1
chr21:17756644:LINC00478:Allele1
chr6:30919878:DPCR1:CADD2
chr10:129160313:DOCK1:CADD1
chr11:35855023::CADD2
chr19:16860558:NWD1:CADD2
chr19:58382542:ZNF814:CADD1
chr4:123609937:IL21-AS1:Allele1
chr16:23711925:ERN2:CADD1
chr4:95587259:PDLIM5:CADD1
chr22:19033747:DGCR2:Allele1
chr20:61472938:TCFL5:CADD2
chr1:35223913:GJB5:CADD2
chr5:65122027:NLN:CADD2
chr19:56466227:NLRP8:CADD2
chr11:209894:RIC8A:CADD1
chr20:61472116:COL9A3:Allele2
chr1:32150411:COL16A1:Allele2
chr19:56466227:NLRP8:Allele2
chr22:19026613:DGCR2:CADD1
chr20:13765507:ESF1:CADD2
chr19:13951830:LOC284454:CADD1
chr22:39631768:PDGFB:CADD2
chr6:112429492:LAMA4:Allele2
chr22:19033747:DGCR2:CADD1
chr10:72288969:PALD1:Allele1
chr7:142239679:TRBV9:CADD1
chr1:170135828:METTL11B:CADD2
chr6:160424760:IGF2R:Allele2
chr1:119606960:WARS2:CADD1
chr11:133815981:IGSF9B:CADD1
chr1:154516477:TDRD10:CADD1
chr10:85898455:GHITM:Allele2
chr2:55872538:PNPT1:CADD2
chr11:112223299:RNU6-44P:Allele1
chr11:2715455:KCNQ1:Allele2
chr16:1894852:MEIOB:Allele1
Variable
0
25
50
75
100
Importance

**B**

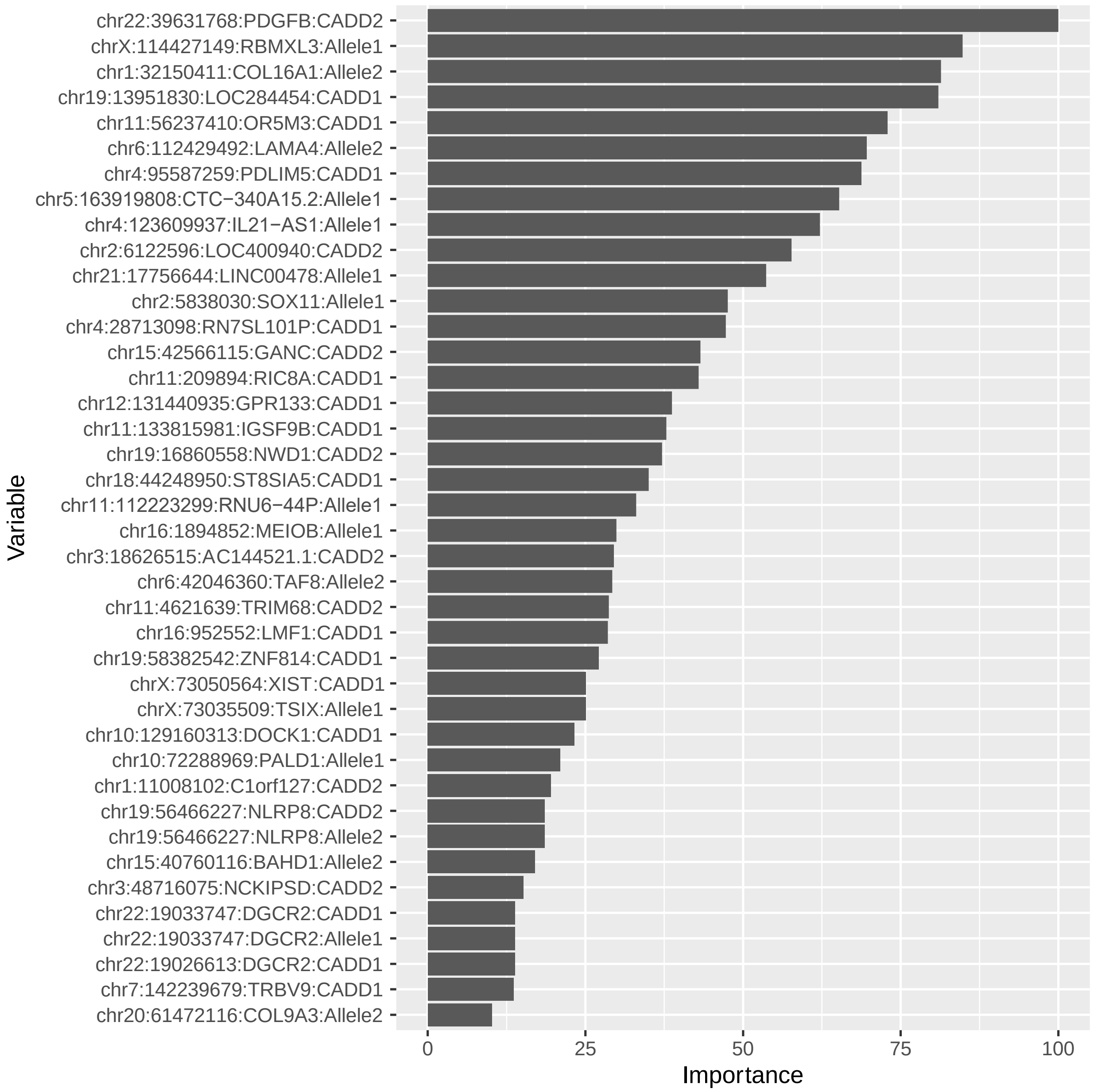

chr22:39631768:PDGFB:CADD2
chrX:114427149:RBMXL3:Allele1
chr1:32150411:COL16A1:Allele2
chr19:13951830:LOC284454:CADD1
chr11:56237410:OR5M3:CADD1
chr6:112429492:LAMA4:Allele2
chr4:95587259:PDLIM5:CADD1
chr5:163919808:CTC−340A15.2:Allele1
chr4:123609937:IL21−AS1:Allele1
chr2:6122596:LOC400940:CADD2
chr21:17756644:LINC00478:Allele1
chr2:5838030:SOX11:Allele1
chr4:28713098:RN7SL101P:CADD1
chr15:42566115:GANC:CADD2
chr11:209894:RIC8A:CADD1
chr12:131440935:GPR133:CADD1
chr11:133815981:IGSF9B:CADD1
chr19:16860558:NWD1:CADD2
chr18:44248950:ST8SIA5:CADD1
chr11:112223299:RNU6−44P:Allele1
chr16:1894852:MEIOB:Allele1
chr3:18626515:AC144521.1:CADD2
chr6:42046360:TAF8:Allele2
chr11:4621639:TRIM68:CADD2
chr16:952552:LMF1:CADD1
chr19:58382542:ZNF814:CADD1
chrX:73050564:XIST:CADD1
chrX:73035509:TSIX:Allele1
chr10:129160313:DOCK1:CADD1
chr10:72288969:PALD1:Allele1
chr1:11008102:C1orf127:CADD2
chr19:56466227:NLRP8:CADD2
chr19:56466227:NLRP8:Allele2
chr15:40760116:BAHD1:Allele2
chr3:48716075:NCKIPSD:CADD2
chr22:19033747:DGCR2:CADD1
chr22:19033747:DGCR2:Allele1
chr22:19026613:DGCR2:CADD1
chr7:142239679:TRBV9:CADD1
chr20:61472116:COL9A3:Allele2
Variable
0
25
50
75
100
Importance

**c**

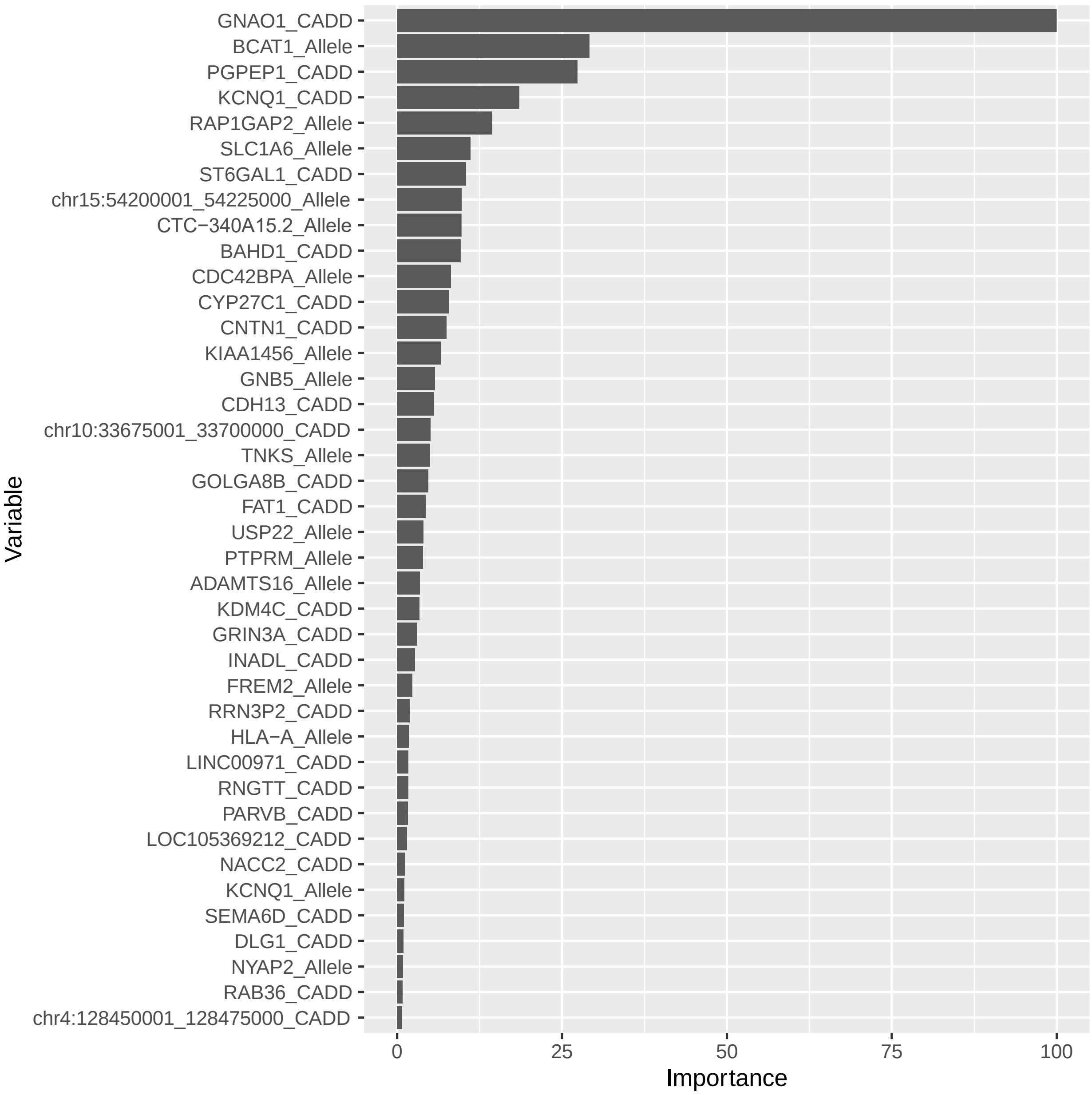

**Supplementary Figure S2**. Permutation-based variable importance top 40 features for top performing dataset and models for the UI cohort loss of smell and/or taste analysis. All features included in the model are assessed. (*A*) Variable importance for the RF model, top performing model for the variant analysis according to the accuracy and brier score metrics. (*B*) Variable importance for the naïve bayes (NB) model, top performing model for the variant analysis according to the accuracy and AUROC curve metrics. (*C*) Variable importance for the XGBTree model, top performing model for the gene analysis.

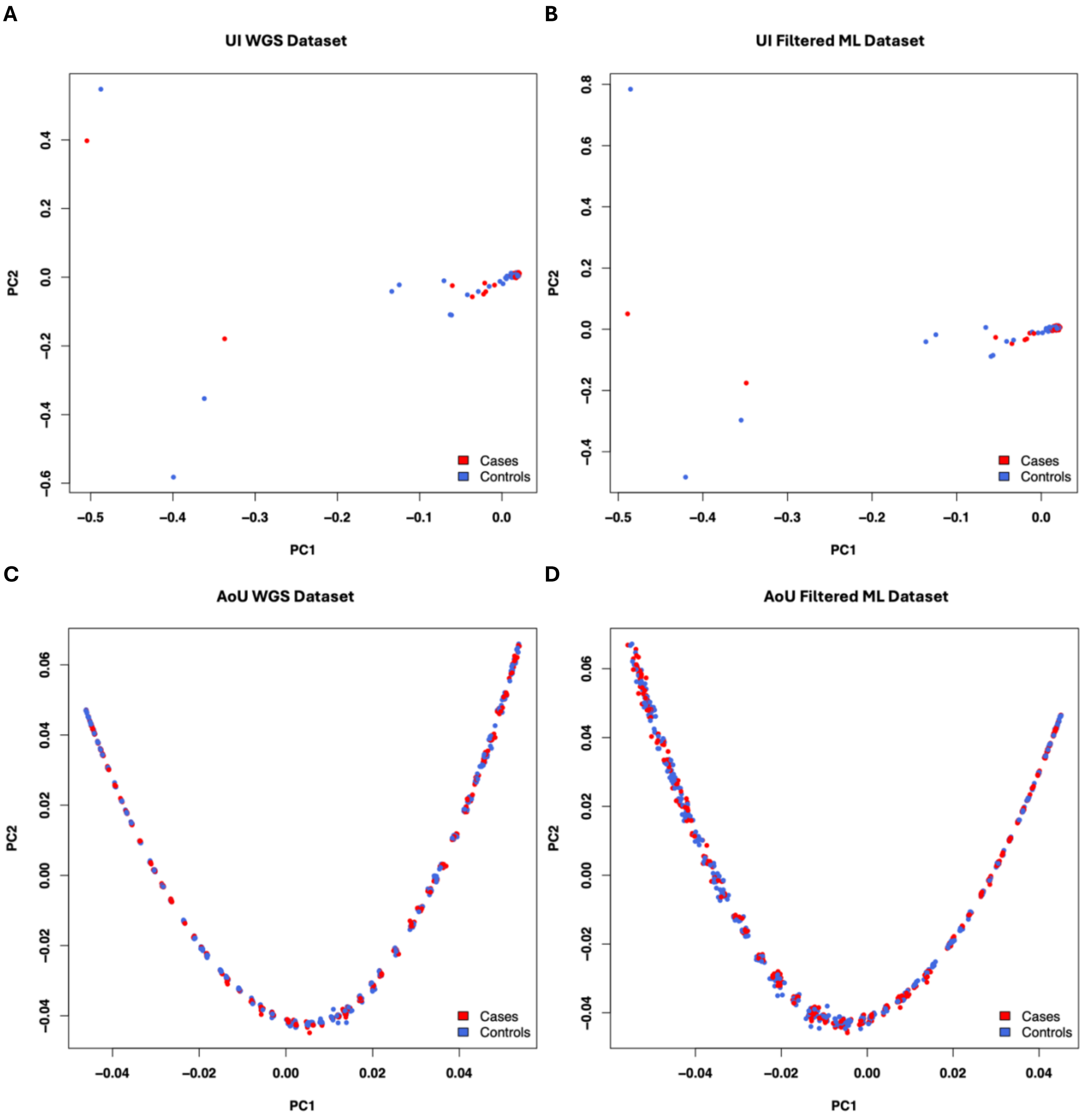
A
UI WGS Dataset
B
UI Filtered ML Dataset
C
AoU WGS Dataset
D
AoU Filtered ML Dataset
PC1
PC2
Cases
Controls

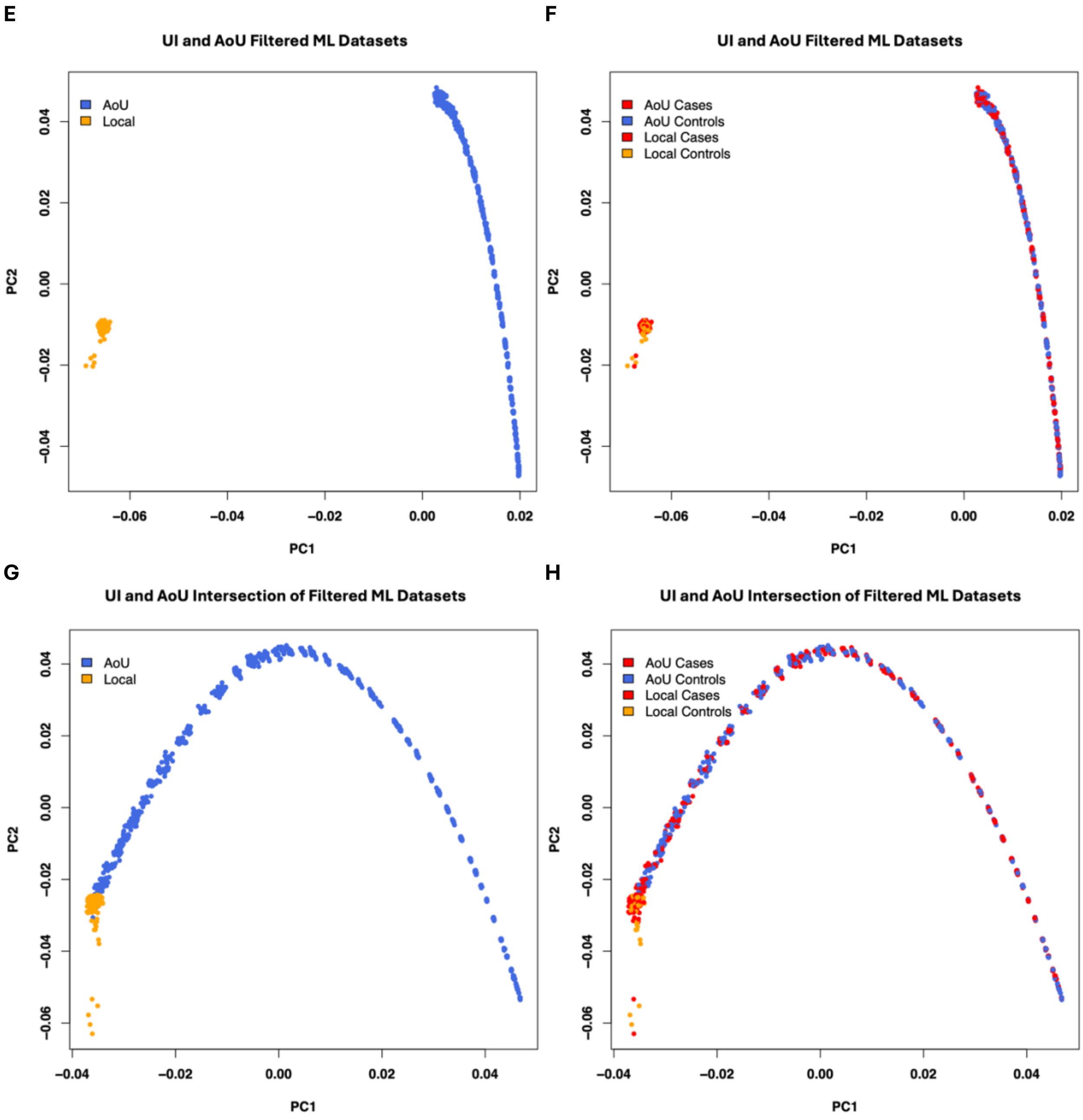
E
UI and AoU Filtered ML Datasets
AoU
Local
PC1
PC2
−0.06
−0.04
−0.02
0.00
0.02
0.04
F
UI and AoU Filtered ML Datasets
AoU Cases
AoU Controls
Local Cases
Local Controls
PC1
PC2
G
UI and AoU Intersection of Filtered ML Datasets
AoU
Local
PC1
PC2
H
UI and AoU Intersection of Filtered ML Datasets
AoU Cases
AoU Controls
Local Cases
Local Controls
PC1
PC2

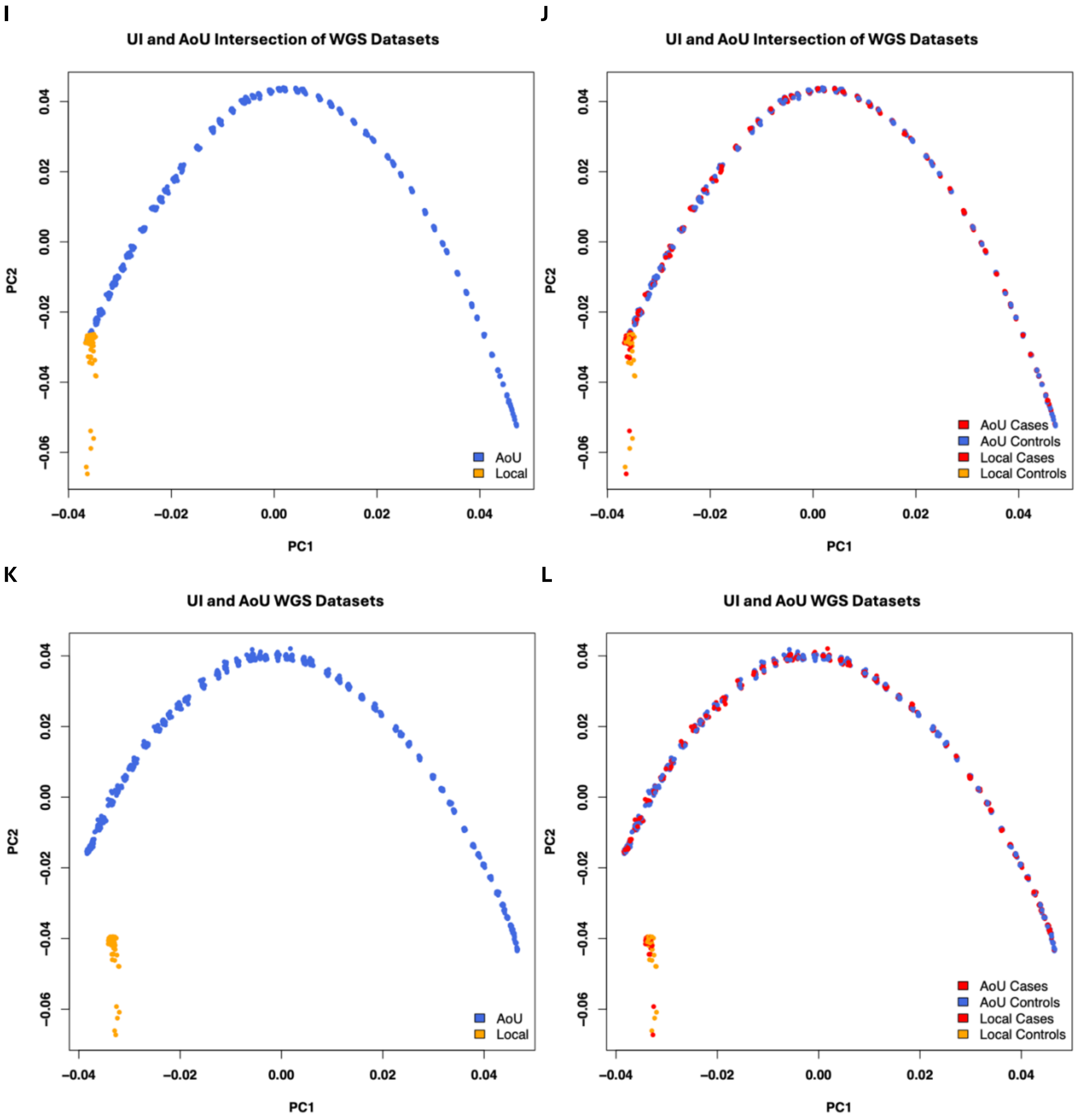

**Supplementary Figure S3.** Comparison of UI and AoU cohort datasets cases and controls and cohorts with principal component analysis, plotting the first two principal components. (*A*) UI whole genome sequencing (WGS) dataset. (*B*) UI filtered ML dataset. (*C*) AoU WGS dataset. (*D*) AoU filtered ML dataset. (*E*) UI and AoU filtered ML datasets. (*F*) UI and AoU filtered ML datasets with cases and controls. (*G*) UI and AoU intersection of filtered ML datasets. (*H*) UI and AoU intersection of filtered ML datasets with cases and controls. (*I*) UI and AoU intersection of WGS datasets. (*J*) UI and AoU intersection of WGS datasets with cases and controls. (*K*) UI and AoU WGS datasets. (*L*) UI and AoU WGS datasets with cases and controls.

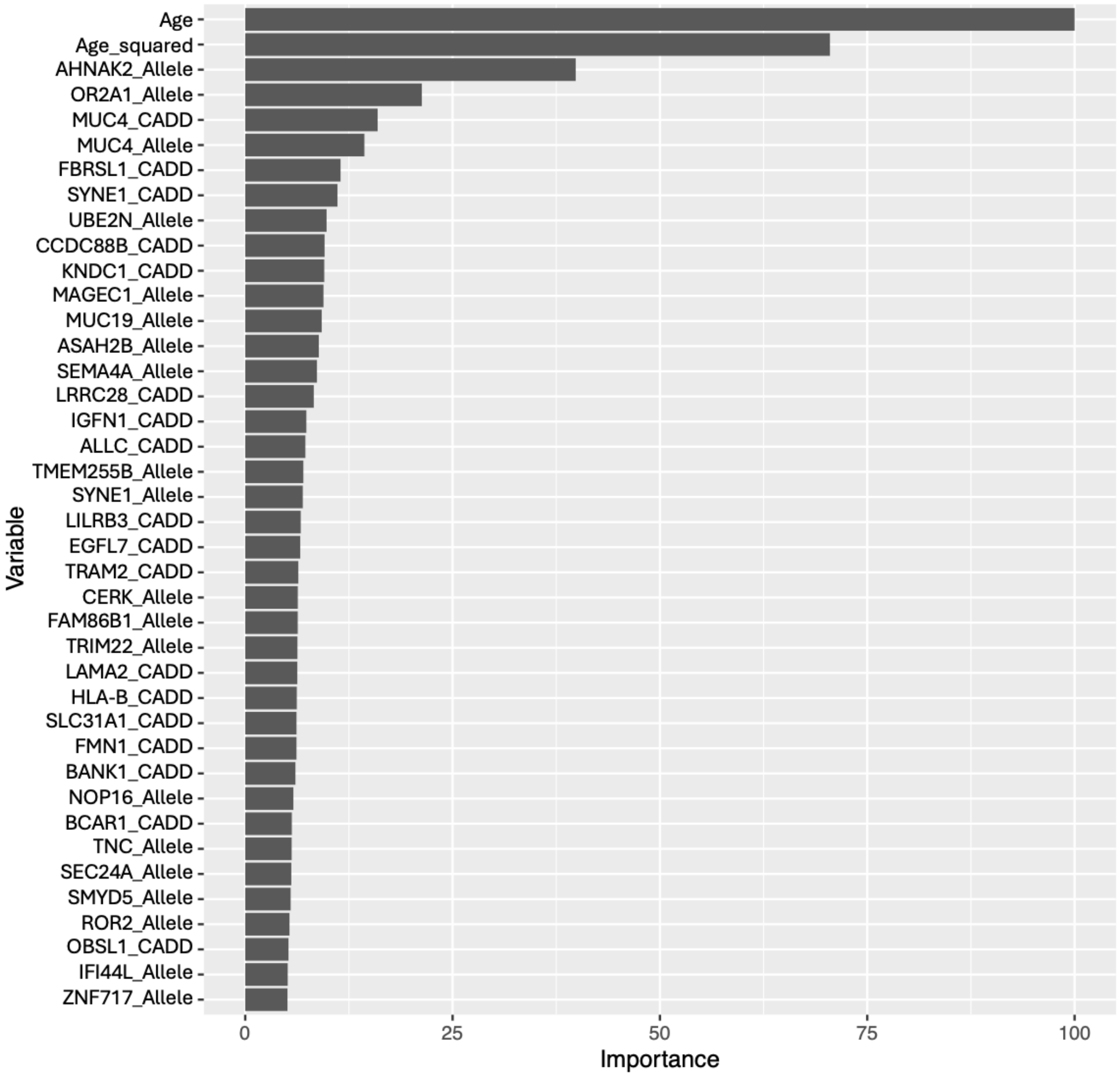

**Supplementary Figure S4.** Permutation-based variable importance top 40 features for top performing dataset and models for the AoU cohort gene analysis. All features included in the model are assessed. Permutations per feature were performed 25 times and performance is assessed based on mean brier score. Variable importance for the RF model, top performing model for the gene analysis.
